# Synthesising environmental, epidemiological, and genetic data to assist decision making for onchocerciasis elimination

**DOI:** 10.1101/2023.02.14.23285937

**Authors:** Himal Shrestha, Karen McCulloch, Rebecca H Chisholm, Samuel Armoo, Francis Vierigh, Neha Sirwani, Katie E Crawford, Mike Osei-Atweneboana, Warwick N Grant, Shannon M Hedtke

## Abstract

**Background:** Population genetics is crucial for understanding the transmission dynamics of diseases like onchocerciasis. Landscape genetics identifies the ecological features that impact genetic variation between sampling sites. Here, we have used a landscape genetics framework to understand the relationship between environmental features and gene flow of the filarial parasite *Onchocerca volvulus* and of its intermediate host and vector, blackflies in the genus *Simulium*. We analysed samples from the ecological transition region separating the savannah and forest ecological regions of Ghana, where the transmission of *O. volvulus* has persisted despite almost half a century of onchocerciasis control efforts.

**Methods:** We generated a baseline microfilarial prevalence map from the point estimates of pre-ivermectin microfilarial prevalence from 47 locations in the study area. We analysed mitochondrial data from 164 parasites and 93 blackflies collected from 15 communities and four breeding sites, respectively. We estimated population genetic diversity and identified correlations with environmental variables. Finally, we compared baseline prevalence maps to movement suitability maps that were based on significant environmental variables.

**Results:** We found that the resistance surfaces derived from elevation (r = 0.793, p = 0.005) and soil moisture (r = 0.507, p = 0.002) were significantly associated with genetic distance between parasite sampling locations. Similarly, for the vector populations, the resistance surfaces derived from soil moisture (r = 0.788, p = 0.0417) and precipitation (r = 0.835, p = 0.0417) were significant. The correlation between the baseline parasite prevalence map and the parasite resistance surface map was stronger than the correlation between baseline prevalence and the vector resistance surface map. The central parts of the transition region which were conducive for both the parasite and the vector gene flow were most strongly associated with high baseline onchocerciasis prevalence.

**Conclusions:** We present a framework for incorporating environmental, genetic, and prevalence data for identifying when ecological conditions are favourable for onchocerciasis transmission between communities. We identified areas with higher suitability for parasite and vector gene flow, which ultimately might help us gain deeper insights into defining transmission zones for onchocerciasis. Furthermore, this framework is translatable to other onchocerciasis endemic areas and to other vector-borne diseases.

## Background

Onchocerciasis is a neglected tropical disease caused by a filarial parasite, *Onchocerca volvulus,* and transmitted by the bites of black flies (*Simulium* spp.). The blackflies have a narrow range of ecological suitability, which leads to spatial heterogeneity in the prevalence and transmission of onchocerciasis [1–4]. The primary tool for onchocerciasis control is mass drug administration with ivermectin (MDAi) with an initial focus on mostly high endemic communities, i.e., there is also spatial heterogeneity in intervention history. Following the success of MDAi in controlling onchocerciasis as a significant public health problem in the majority of areas, almost all countries have switched their target from control to elimination. However, the target of onchocerciasis elimination with MDAi is impeded by some persistent onchocerciasis transmission foci despite decades of intervention [5–7].

Understanding the persistence of disease transmission requires spatial heterogeneity to be considered because of the risk that movement of infective vectors, and thus parasites, from the areas with different endemicity and MDAi history can re-initiate disease in areas where transmission of *O. volvulus* is thought to be eliminated. For instance, the migration of the parasites via infected humans has been linked to recrudescence in previously eliminated foci of Burkina Faso [8–10]. Similarly, the failure to achieve the elimination of onchocerciasis in West Africa with the onchocerciasis control program (OCP) was attributed to rapid insecticide resistance due to high vector gene flow and, thus, the spread of insecticide resistance alleles [11–14]. However, disease control programs have historically focused on government administrative units as the unit of intervention, which has led to a situation where treatment decisions are being made without much consideration of host- or vector-mediated movement of the parasites and, thus, the transmission zones.

The geographical unit in which parasite transmission occurs via locally breeding vectors is termed as a transmission zone [15]. Transmission zones form the biological basis of intervention units, and thus, a clear understanding of transmission zones and means to define their boundaries are crucial to ensure that the interventions are coordinated at the correct geographic scale. Onchocerciasis prevalence is high in the poorest of the poor nations of the world [12, 16]. Therefore, the limited resources available in these areas must be judiciously allocated to the most essential areas to achieve the elimination of onchocerciasis transmission. The way forward to achieving the elimination goal is to align intervention units as closely as possible to the natural transmission zones. However, delineating transmission zones is a challenging task, and several tools have been deployed so far to understand transmission zones.

We can gain some insights into the transmission zones based on prevalence mapping, where point prevalence data are interpolated spatially [4, 17]. However, this is a static map and ignores the ‘innate’ connectivity between locations mediated by the movement of the human host and the vectors. Population genetics has been used to infer the movement of pathogens, whereby pathogen movement can be measured indirectly by the genetic relatedness of parasites across locations [18–28]. The dispersal, and thus gene flow, of parasites and vectors, are subject to influence by the environmental features of the landscape. Therefore, population genetics should be combined with spatial information and environmental data in order to provide a better picture of the transmission processes. This combination of spatial information, environmental data and population genetics is termed landscape genetics.

Landscape genetics explicitly quantifies the effects of landscape on evolutionary processes such as gene-flow, drift, and selection [29, 30]. Spatial information can be added in the form of sampling location geographic coordinates and remote sensing satellite images of different environmental and climate variables such as elevation, slope, distance to the water bodies etc. There are then several steps required in order to use landscape genetics to infer transmission zones. First, the degree of genetic differentiation between sampling locations for parasites and/or vectors is measured. Second, the extent of correlation between a range of environmental variables and the measures of genetic differentiation estimated in step one is determined [31]. Third, the most important environmental variables identified in step two are converted to resistance surface maps, which quantify the barriers to the gene flow of the study population in a pixel-level landscape map and are a proxy for the movement suitability of an organism in that particular landscape, i.e., high resistance implies low gene flow/mobility and low resistance implies high gene flow/mobility [32, 33]. Resistance maps can be used to simulate the pattern of gene flow of the parasites and the vectors, giving insights into the predicted corridors of movement and, thus, the likelihood of transmission between locations [34, 35].

We have implemented this technique in the ecological transition region of Ghana, an onchocerciasis hotspot of concern. Despite half a century of interventions, *O. volvulus* transmission still persists in some communities [5,36,37], and there are also reports of suboptimal response (SOR) of infections to treatment with ivermectin [38–40]. A recent population genetic analysis by Crawford et al. [25], suggested a genetically homogeneous parasite populations in this area with the absence of isolation-by-distance, i.e., genetic connectivity of the parasite population not limited by the geographic distance between the population. This suggests cross-transmission of *O. volvulus* between communities, which may be contributing to the persistence of onchocerciasis transmission. With the hypothesis that the genetic connectivity is influenced by environmental factors, we used a landscape genetics framework to understand the spatial patterns of transmission in the ecological transition region of Ghana.

We have combined environmental data with the parasite genetic data (and have included additional vector genetic data from the ecological transition regions) with the objectives of: (i) determining the ecological factors affecting the spatial variation in the parasite and the vector population genetic estimates and; (ii) inferring the patterns and routes of gene-flow, and thus the likely transmission, for the parasite and the vector populations. We have identified key environmental variables that influence the population genetic structure of the parasite and the vector population and generated gene flow maps for the parasite and the vector population from the ecological resistance surface maps. This allowed us to identify potential corridors of parasite and vector movement between the sampling communities, which provides an evidence base for spatial delineation of transmission zones. Further, we have compared the movement suitability maps with the baseline microfilarial (mf) prevalence maps and discussed the immediate implications of the approach developed to aid elimination goals.

## Methods

### Sampling locations

The study area is a west-east transect in the ecological “transition zone” of Ghana: an area that includes the savannah ecotype in the north and the forest ecotype in the south [41–43], with the Lake Volta bisecting the eastern parts of the transition zone, and the Bui National Park in the west (Figure 1). We chose this area for the study as there is ongoing persistence of *O. volvulus* transmission despite decades of control efforts [36,40,44]. The elevation ranges from 70–525 m above sea level, and mean annual temperature and precipitation range from 24– 29°C and 1077–1355 mm, respectively [45, 46].

**Figure 1.**
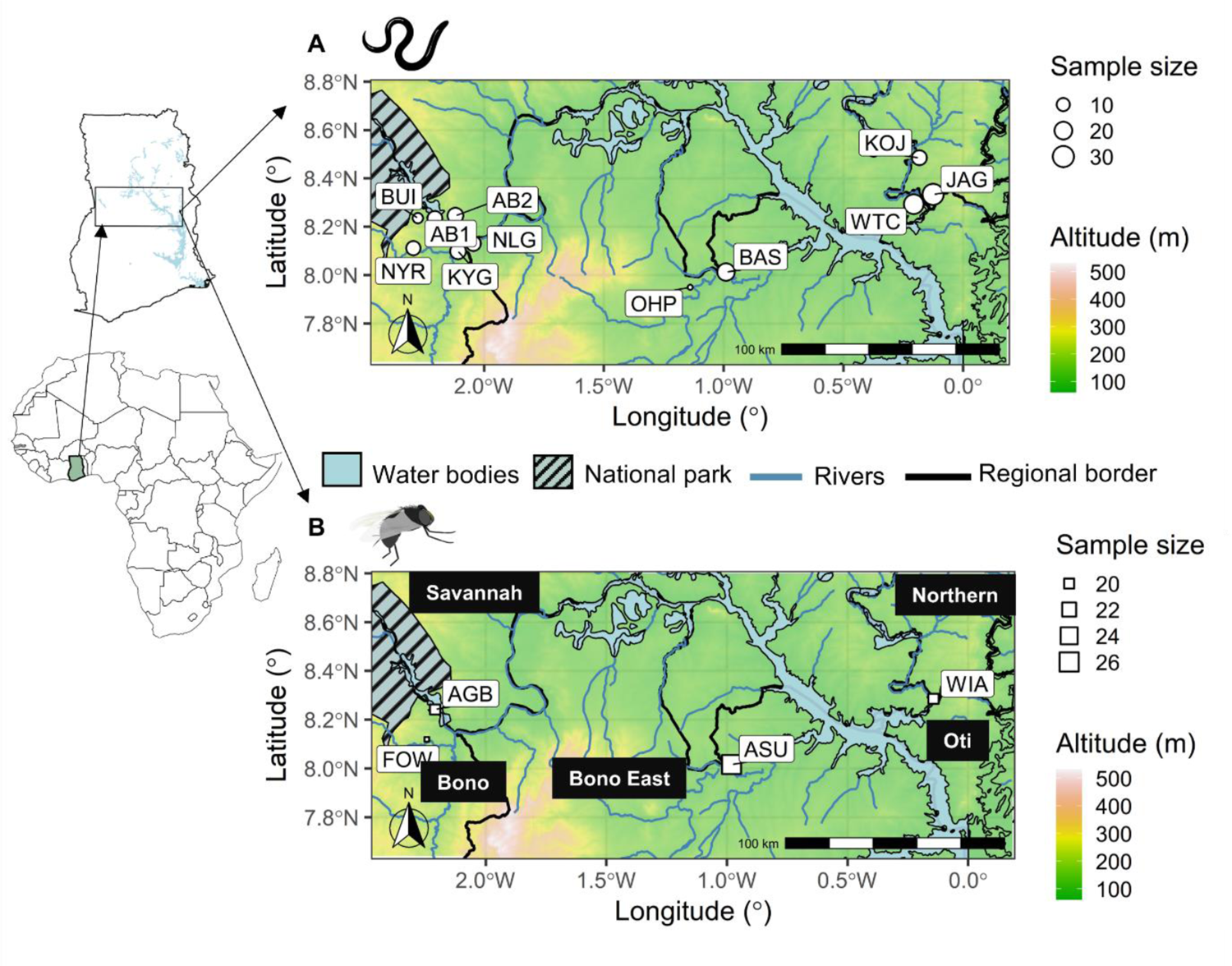
The spatial context of the sampling locations of the *Onchocerca volvulus* and *Simulium damnosum* in the transition region of Ghana. Geographic coordinates are represented as the circle for parasites (**A**) and square for vectors (**B**), and their sizes correspond to the number of samples from the respective locations. The legend for the size is provided to the left of each figure. The communities are represented with community codes. The river lines and the government administrative borders are shown along with the water body (Lake Volta) and the Bui national park. The inset map shows the map of Africa and Ghana with the bounding box for our study area. More information about sampling locations and the number of samples are present in Additional file Table S3.

The sampling locations belonged to four different government administrative regions, viz., Bono, Bono East, Savannah, and the Northern regions (Figure 1; Additional file Table S3). Variant calls based on mitochondrial genome data from 164 female *O. volvulus* samples that had been isolated from 97 people from 15 communities, primarily in 2010–2012 (n = 107) and in 2006 (n = 34) and 2013 (n = 23), were obtained from Crawford et al. [25]. Ethics approvals for sampling parasites from people are reported in Crawford et al. [25]. Four communities from each of the four regions, viz., Bono, Bono East, Savannah and Northern region, were chosen for the sequencing of vector samples which were collected in 2013–2015. A total of 93 *S. damnosum* samples collected in 2013 (n = 73) and 2015 (n = 20) by human landing catch were selected from four communities.

A bounding box formed based on the convex hull boundary (a boundary with a set of convex curves enclosing the sampling locations) around the sampling locations was used for the geospatial analysis. The dimension for the bounding box was 293.68×129.38 km (an area of 37,995.59 km^2^). Geographic coordinates for all the communities were used to calculate the pairwise geographic distance between the communities (Additional file Table S3). We aggregated data from communities close to each other (less than 5 km) and used the centroid of the geospatial coordinates of the communities in close proximity for the merged communities. This brought the number of parasite sampling locations down to 11 but increased the sample size per community (Figure 1).

### Sequencing and variant calling

Details on the genetic data generation and the parasite samples are available in Crawford et al. [25]. In brief, DNA was extracted from adult female *O. volvulus* from nodules using the Dneasy^®^ Tissue Kit (Qiagen, Hilden, Germany) following the manufacturer’s instructions. Sequence libraries were generated based on either genomic DNA extracts or on amplicons targeting the mitochondrial genome and sequenced using Illumina MiSeq or HiSeq sequencing platforms. Trimmed sequence reads were mapped to the *O. volvulus* (NC_001861) mitochondrial reference genome and variants called using *GATK UnifiedGenotyper* [47]. These data were submitted to the NCBI Short Read Archive under project PRJNA560089 [48].

For *S. damnosum*, the body of each fly was dissected and homogenised using a pestle. Extractions of total DNA were performed using the Isolate II Genomic DNA kit, following the manufacturer’s instructions (Bioline, London, United Kingdom). Sequencing libraries were constructed and indexed using the Illumina DNA Prep tagmentation kit following the manufacturer’s instructions (Illumina, San Diego, California, USA). Libraries were pooled and sequenced on one lane of a NovaSeq SP, 300 cycles (resulting in 150-bp paired-end reads) at the Australian Genome Research Facility (Melbourne, Victoria, Australia) (Additional file Table S1).

Sequenced reads were trimmed for quality and to remove adapter contamination using *trimmomatic* v.0.32 and keeping only those pairs where both pairs were >125 bp [49]. To assemble the genome, three flies with the largest number of paired reads were mapped using *bwa* v. 0.7.17 [50] to available *Simulium* spp. Complete or nearly complete mitochondrial genomes downloaded from NCBI (*Simulium variegatum*, NC_033348; *Simulium noelleri*, NC_050320; *Simulium quinquestriatum*, MK281358; *Simulium ornatum*, MT410845; *Simulium maculatum*, NC_040120; *Simulium aureohirtum*, NC_029753; *Simulium petricolum*, MT671497; *Simulium equinum*, MT920425; *Simulium angustipes*, MT628576; *Simulium lundstromi*, MT628562). Those reads that mapped to any *Simulium* genome were extracted and converted to fastq using *samtools* v.1.9 [51], and these were used to produce a preliminary assembly using *spades* v. 3.11.1 [52] and *velvetoptimiser* v. 2.2.5 [53, 54]. These drafts were then improved using *pilon* v.1.23 [55]. Assemblies from the two different programs were aligned in Mesquite v.3.61 [56], and the consensus—defined as bases that were observed in both assemblies—was taken to produce a single consensus reference genome (i.e., the consensus from two variant callers from one blackfly) for variant calling. Because mitochondrial genomes are circular, and thus the starting point for different linear assemblies differed, the assembly for each fly was oriented so that it began with tRNA-Ile to be consistent with *S. variegatum* (NC_033348; [57]). The “AT-rich region” was variable in inferred length and sequence between different assemblers, different individual blackflies, and different species, and were difficult to align. Thus, this AT-rich, variable-length region was excluded. All raw reads and assembled sequences were submitted to the European Nucleotide Archive (ENA) at EMBL-EBI under accession number PRJEB57094.

Variants were filtered to retain only those calls at positions with a minimum quality score of 30 and a minimum depth of 20 using *vcftools* v.01.13 [50,58,59]. Individuals with more than 75% missing data were excluded from the analysis. Variants were normalised using *bcftools* v.1.2. To ensure consistency between variant formatting, allelic primitives were called using the function *vcfallelicprimitives* implemented in *vcflib* [60]. The intersection of the two variant callers was then identified using *bcftools* v.1.2 [61]. For both parasite and vector data, we filtered the variants to remove indels, missing regions, and non-biallelic sites using *vcftools* v.01.13 [58]. The resulting dataset comprised 189 SNP loci for 164 individual *O. volvulus* and 632 SNP loci for 93 individual *S. damnosum*.

### Prevalence data

Pre-MDAi prevalence data for communities that fell within the study area bounding box and were based on observation of mf in a skin biopsy via microscopy were obtained from the Expanded Special Project for Elimination of Neglected Tropical Diseases (ESPEN) database [62]. The prevalence data collected for mapping, i.e., prior to MDAi was used. Duplicate observations were removed, and observations from the same geographic coordinates at different years were aggregated to calculate the average prevalence. There were 47 unique locations with prevalence data collected from 1976 to 2004 that fell within the study area used for the geospatial mapping of the baseline prevalence.

### Environmental data

We compiled different continuous environmental rasters which might be ecologically relevant to the onchocerciasis distribution based on the published literature, field experiments on blackflies [63, 64] and ecological factors identified with previous geospatial modelling studies [2,17,65–67]. These environmental variables included distance to the nearest river, soil moisture, elevation, slope, temperature, and precipitation [2,65,67]. In addition, the dispersal capacity of the *Simulium* vector is dependent on the vegetation type and time of the year [68]. Therefore, we included vegetation and seasonality-related variables in our analysis. In addition to environmental variables, we also included some sociodemographic aspects of the study area—for example, the human population density to consider the availability of human hosts for disease transmission. We used the environmental variables corresponding to the year when the samples were collected for fitting the models to account for the differences in the time of sampling. For prevalence data, environmental variables before 2001 were used, and similarly, for the *O. volvulus* and *S. damnosum*, environmental variables from 2010–2012 and 2013– 2015 were used respectively, as per the data availability. Our starting set of environmental and socio-economic datasets consisted of 32 continuous environmental rasters at a spatial resolution of 1 km from publicly available repositories via Earth Engine (Additional file Table S2) [69].

These variables were divided into six groups, viz., temperature, precipitation, topography, vegetation indices, hydrological and sociodemographic variables. We extracted the values for each sample location using the *raster* package in R v. 4.1.0 [70, 71]. For testing the association of the landscape factors to the genetic differentiation or gene flow between the populations, a pairwise comparison of environmental characteristics between sampling locations is crucial [29, 72]. Thus, we calculated the average of the values encountered by a pairwise straight path between each sampling site to account for the features in adjacent areas around sampling sites for all the environmental and sociodemographic variables. We generated a pairwise correlation matrix for all 32 variables to identify variables that are highly correlated with prevalence (Additional file Figure S2, S4). We included only those variables where Pearson’s correlation coefficient between the ecological variable(s) and prevalence was less than < |0.6| within each group of variables [72]. Further, we performed principal component analysis (PCA) to identify the variables that contributed most to the variance among the group of correlated variables (Additional file Figure S1, S3) [73]. For any given group of correlated variables, we selected the variable with the highest contribution score to the total variance in PCA analysis and the ease of interpretability of the variables. The environmental variables selected for the parasite sampling locations were also used for vector landscape genetics for easier comparison between the vector and the parasite landscape genetics.

### Prevalence mapping

The mean of the posterior prevalence was obtained from the pre-MDAi mf prevalence data using the Bayesian approach with Integrated Nested Laplace Approximation (INLA) [74, 75]. The number of positive cases out of the total number of people tested in a location was assumed to follow a binomial distribution. The prevalence was modelled with different environmental variables and a spatial random effect with a zero-mean Gaussian process following a Matérn covariance function. The Matérn field is represented with a finite element mesh formed of triangles around the sampling locations and adding vertices over the prediction region. Multiple triangulation meshes with different parameters for cut-off and length of triangles inside and outside the boundary were tested for the model fit and computational cost (Additional file Figure S5). We created a triangulation mesh with a 3 km cut-off; the maximum length of triangles inside and outside the boundary was set to 10 km and 100 km, respectively. Finally, we fitted the model and assessed the relationship of environmental variables with the prevalence data. The details of fitting a spatial model to the prevalence data for geospatial mapping are available in [2]. The prediction of the posterior prevalence was made at a 2 km resolution considering the high computational cost of prediction on a lower resolution.

### Population genetic analysis

For the parasite and the vector samples, we carried out unsupervised *k*-means clustering analysis using the *adegenet* v. 2.1.6 package [76]. We inferred the optimal number of *k* (groups) for the population using unsupervised *k-*means clustering with the Bayesian Information Criterion (BIC). The vector results were consistent with the results of a haplotype network analysis using *PopART* [77] that identified outlier blackflies separated largely from the cluster of other samples. Given the taxonomic uncertainty of the species composition of the *S. Damnosum* complex, these outliers could not be assigned confidently as members of the same interbreeding population that we believe comprised the bulk of the black flies in the sample and were therefore excluded from the analysis. Then, we carried out a Discriminant Analysis of the Principal Components (DAPC) using communities as populations. DAPC is sensitive to the number of principal components retained. Therefore, we performed stratified cross-validated DAPC by varying the number of principal components using *xvalDapc* function in the *adegenet* v. 2.1.6 package. We calculated the membership probability of each sample, communities, and the posterior correct assignment probability for the communities. We calculated summary statistics for the genetic data, i.e., number of alleles, observed gene diversity, and the pairwise measure of genetic differentiation (F_st_) between sampling locations using the *Hierfstat* v. 0.5.11 package [78]. Similarly, mean allelic richness and number of haplotypes were calculated using *PopGenReport* v. 3.0.4 and *haplotypes* v. 1.1.2 package, respectively [79, 80]. The pairwise F_st_ matrix was adjusted for finite populations by linearising it with the equation F_st_/(1 − F_st_) as suggested by [73,81,82].

### Landscape genetic analysis

Landscape genetics analysis helps us understand how landscape features influence the spatial distribution of genetic variation. The simplest starting model is the isolation-by-distance model, where we test if there is a correlation between the pairwise genetic distance and the pairwise straight-path geographic distance between the sampling sites [30,83,84]. The geographic distance was calculated as the pairwise Euclidean distance between the geographic coordinates of the sampling sites using the *graph4lg* v. 1.6.0 package [85]. Geographic coordinates were converted to the Universal Transverse Mercator projection, a two-dimensional cartesian coordinate referencing system that is accurate when performing distance-related operations on spatial objects [86]. The coordinate referencing system used in our analysis for all the spatial objects was: `epsg-32630 (+proj=utm +zone=30+datum=WGS84 +units=m +no_defs)`. The pairwise linearised genetic differentiation between sites was considered a genetic distance. We performed Mantel tests between the geographic distance and the genetic distance matrix with the *vegan* v. 2.6.2 package, and the significance of the correlation was calculated based on 10000 permutations [87].

### Resistance surface maps

In addition to geographic distances, we calculated ecological cost distances to assess the effect of intervening landscape features between the sampling sites on spatial genetic variation [31, 88]. The ecological cost distances were calculated based on “resistance surface” maps. The values in each pixel of a resistance surface map reflect the extent to which the landscape feature on that pixel impedes or facilitates the movement or connectivity of the populations of interest between different locations [33, 35]. We used *Circuitscape* implemented in Julia v. 1.6.1 to calculate the circuit distance, a proxy for the ecological cost distances, to generate connectivity maps and identify corridors for movement in the landscape [89].

The resistance surface maps were generated from the environmental variables using a search and optimisation method, where transformation parameters were explored to maximise the association between the pairwise genetic distance and the ecological cost distance using *ResistanceGA* v. 4.1.46 package [33]. The package uses a genetic algorithm to optimise resistance surface parameters and offers eight transformations of ricker and monomolecular functions to a continuous surface. The following equations give the ricker and monomolecular transformation function:

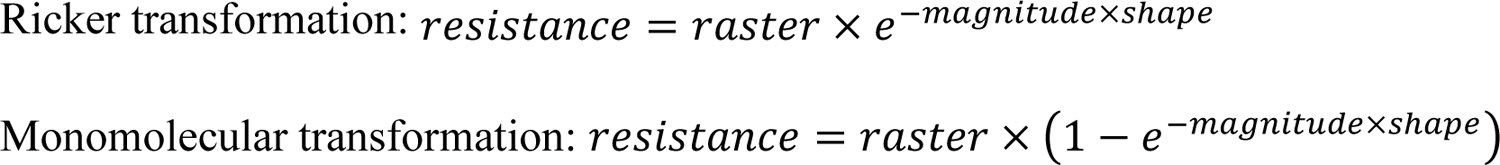

The algorithm searches for the best combination of a transformation function, magnitude, and shape parameter. It provides a framework for optimising resistance surfaces from an environmental raster surface without any prior assumptions about the contribution of those surfaces on the resistance [33] and, therefore, provides an unbiased representation of the resistance surface based on genetic data.

The environmental variables selected for landscape genetic analysis were used to optimise the resistance surface maps. Linearised pairwise F_st_ genetic distance between sampling locations was used as the response parameter. The cost distance calculated from the transformed resistance surfaces was used as a predictor to find the best model that explains the genetic distance. A linear mixed-effects model with a maximum likelihood population effect (MLPE) was fitted to the data [90, 91]. We optimised single surfaces of environmental variables and used the log-likelihood as the objective function for the MLPE model. Four replicates of 1000 iterations each were run with the optimisation set to stop after 50 generations of no improvement. We set the maximum allowable resistance value to 100 during the optimisation process for easier rescaling and comparison of the resistance values of different environmental variables.

Each replicate of the resistance surface obtained via the optimisation process was tested using the circuit distance matrix obtained from those resistance surfaces. We used the partial Mantel test to assess the correlation between the genetic distance matrix and the pairwise circuit distance matrix accounting for the geographical distance matrix. The partial Mantel test is used frequently in landscape genetics analyses but has high type I error rates with spurious correlations [92]. Therefore, we used mixed matrix regression with randomisation (MMRR) as a confirmatory test. The MMRR was performed using the *lgMMRR* function in the *PopGenReport* v. 3.0.4 package based on Wang’s (2013) method. The MMRR also gives us the effect of the resistance surface on the genetic differentiation accounting for the geographic distances. To avoid spurious correlations, we took a conservative approach, and the resistance surfaces were deemed significantly associated with the genetic distance only if both the partial mantel and MMRR tests were statistically significant [73, 94]. Significance for both the partial Mantel and MMRR were assessed based on 10,000 permutations.

### Composite resistance surface maps

As landscape features and environmental gradients do not exist in isolation, the environmental resistance surfaces significantly associated with the genetic distance matrix were manually combined to form a composite resistance surface map. They were rescaled from 0 to 1, where the maximum resistance value among all the significant surfaces was considered as 1, preserving the relative contribution of each optimised surface to the composite resistance map. The composite resistance map was obtained by multiplying the rescaled significant resistance surfaces described in Schwabi et al. [31]. The composite resistance surfaces were used for connectivity mapping and identifying corridors of movement via Circuitscape v. 5.10.2 [34, 89]. A bivariate map of posterior mean prevalence was plotted with composite resistance surface maps to visualise areas of varying prevalence and resistance. Correlation coefficients between the mean prevalence map and both the vector and parasite composite resistance surface maps were calculated. We also generated bivariate moving window correlation measures, their significance, and Moran’s-I measure of spatial autocorrelation to measure the correlation between two spatial processes [95].

## Results

### Prevalence mapping

For the analysis of the prevalence data, the land surface temperature at night, temperature seasonality, minimum temperature of the coldest month, soil moisture, annual precipitation, slope, distance to the nearest river and prevalence of improved housing were selected. mf prevalence data ranged from 0.65% to 82.95% with a mean of 29.01% (± 19.31% SD). Most of the data were from the western and south-central parts of the study area, with only five data points from the eastern parts (Figure 2a). The geostatistical interpolated map of baseline mf prevalence based on environmental data shows that the prevalence is higher, particularly in the south-central, central, and eastern areas of the transition Ghana (Figure 2c). The overall predicted prevalence is relatively low in the western areas of transition Ghana with scattered areas of high prevalence. As expected, the uncertainty map shows that the uncertainty was relatively lower in the actual sampling locations with varying levels of uncertainties in the interpolated areas (Figure 2d). Based on the regression coefficients, the soil moisture (mean coefficient: 0.043, 95% BCI: 0.004–0.084) and slope (mean coefficient: 2.126, 95% BCI: 0.032–4.338) had a significant positive association with the mf prevalence while the temperature seasonality (mean coefficient: −0.022, 95% BCI: −0.044–-0.001) had a significant negative association with the mf prevalence (Additional file Table S4). The spatial range of the mf prevalence map was estimated to be 4.4 km (95% BCI: 1.67–7.88 km).

**Figure 2.**
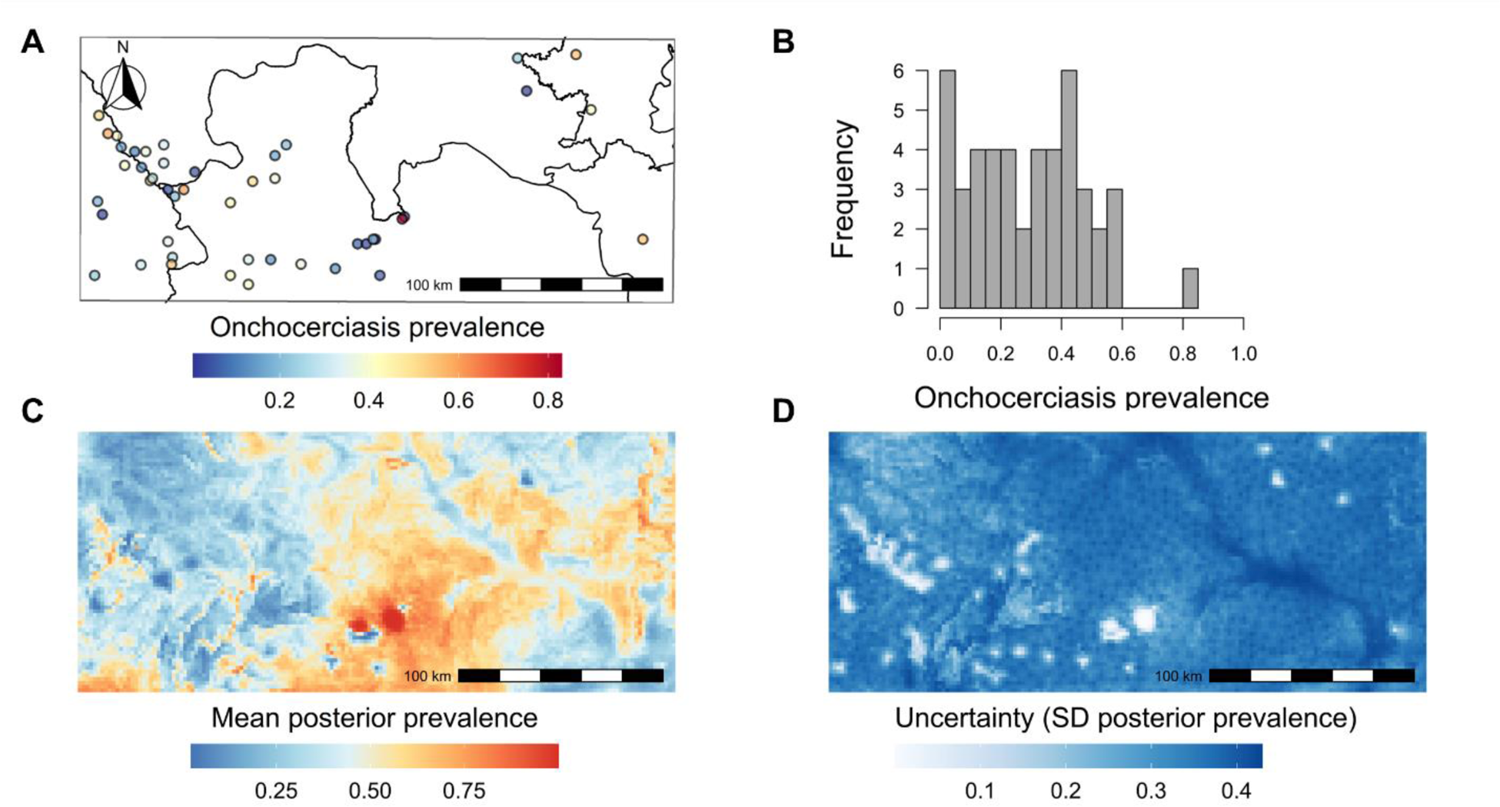
Mapping baseline prevalence of *Onchocerca volvulus* infection in the transition region of Ghana. Pre-MDAi point microfilarial prevalence data (*n* = 46) (**A**), where circles represent sampling locations and the colours of the filled circles represent prevalence according to the heat bar below the figure. The solid line indicates the regional boundary. (**B**) shows the histogram of the pre-MDAi mf prevalence data. The model predicted estimate of the baseline prevalence of *O. volvulus* infection (**C**) in the transition region of Ghana and the uncertainty, i.e., the standard deviation (SD) of the posterior prevalence (**D**) is shown in the bottom row.

### Population genetic analysis

We carried out unsupervised *k*-means clustering analysis and visualised the haplotype network for both the parasite and the vector mitochondrial data separately to observe if there were any inherent clusters and if there were any outlier samples. We chose the minimum number of principal components that explained the highest cumulative variance. The number of principal components retained for the clustering analysis of the parasite and the vector was 80 and 45, respectively. We chose the number of optimal clusters based on the BIC scores, i.e., *k* = 8 for the parasite data and *k* = 12 for the vector data, as the decline in BIC saturated beyond these values (Additional file Figure S6). The clustering and haplotype network analysis on the *Simulium* data indicated the presence of outliers (groups 6 and 10; Additional file Figure S7) which were removed in the downstream analysis. For the parasite samples, the number of alleles and the number of haplotypes corresponded to the sample size of the population, while the mean allelic richness and the gene diversity correlated with each other (Additional file Table S3). The number of principal components was optimised as 72 and 40, respectively. DAPC for the parasite genetic showed overlap between the clusters of the communities, except for a few communities like OHP and NLG (Figure 3). The average percentage of the correct assignment for parasites was 71.21% (±11.45% SD), which would generally be considered relatively poor. For vectors, DAPC also showed low overlap between clusters of the communities and an average % correct assignment of 74.03% (±8.36% SD). The mean percentage reassignment was not significantly different (*p* = 0.62) between parasites and vectors, i.e., DAPC showed that the spatial distribution of parasite and vector genetic variation was similar.

**Figure 3.**
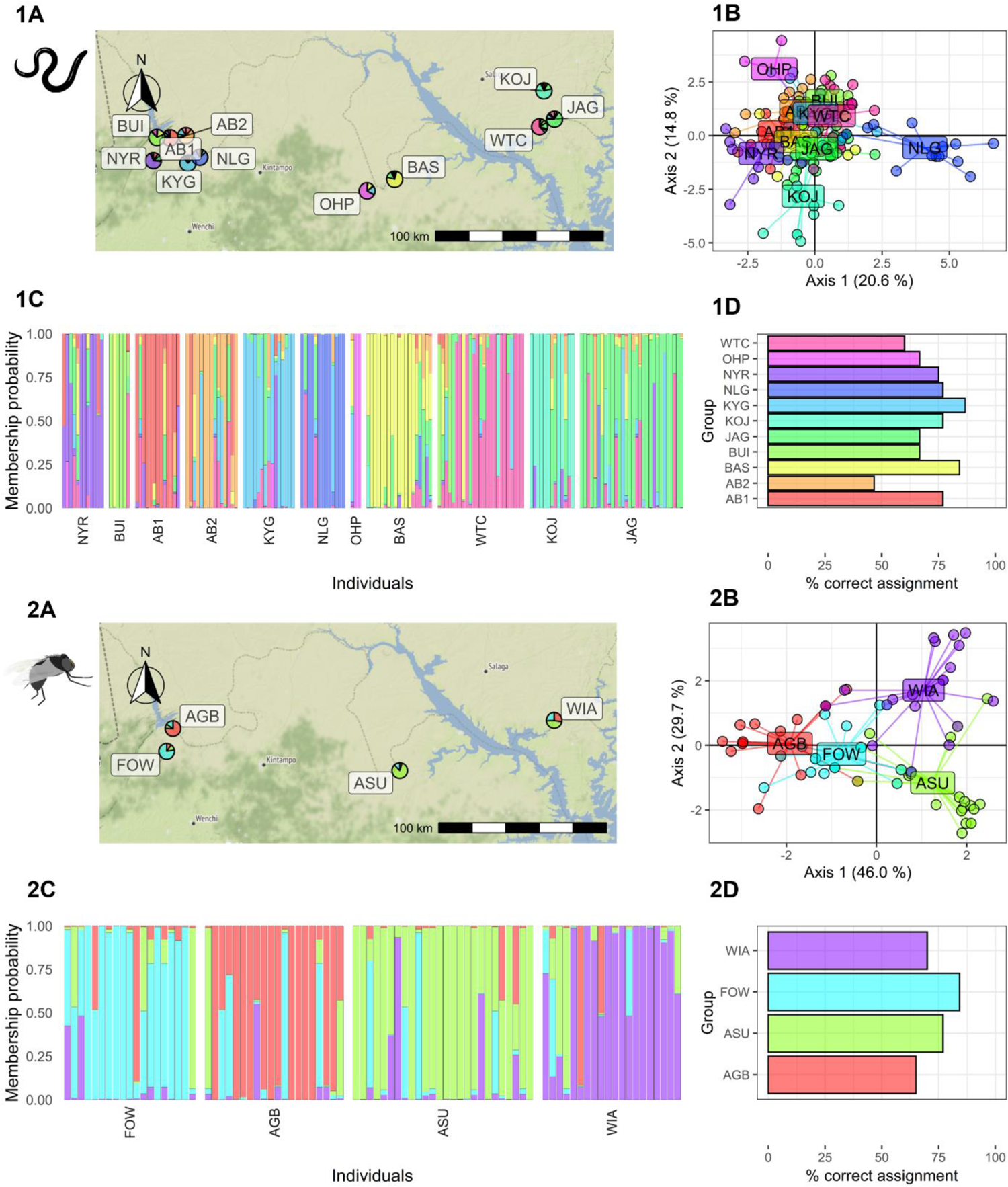
Discriminant analysis of the principal components (DAPC) analysis for the parasite and the vectors sampled from 11 and 4 communities respectively in the transition region of Ghana. The pie chart on the map (1A, 2A) indicates the community level of membership probability. The DAPC analysis shows the community clusters (1B, 2B) and the individual level membership probability (1C, 2C) with each block representing communities. The percentage of the samples assigned correctly to their respective communities is shown for both the parasites (1D) and the vectors (2D). The community codes are presented in Additional file Table S3.

### Landscape genetic analysis Isolation-by-distance

The Euclidean distance matrix between sample locations and the matrix of linearised pairwise F_st_ was used to test whether the parasites and vector population structure conformed to an isolation-by-distance model, in which the degree of genetic differentiation is correlated positively with geographic distance between sampling locations [84]. The Euclidean geographic distance between locations ranged from 2.2 km to 240.39 km. For the parasite sampling locations, six communities were less than 5km apart and were merged into two communities. The geographic distance for the parasites averaged 117.73 km (±11.50 SE; range: 7.86–240.43 km), and the genetic distance averaged 0.11 (±0.009 SE; range: 0.041– 0.286). Similarly, for the vectors, the geographic distance for the parasites averaged 141.40 km (±33.61 SE), and the genetic distance averaged 0.056 (±0.007 SE; range: 0.04–0.084). The Mantel test indicated a poor correlation between the genetic distance and the geographic distance for both the parasite (Mantel’s r = −0.052; p = 0.543) and the vector data (Mantel’s r = −0.039; p = 0.583) (Figure 4).

**Figure 4.**
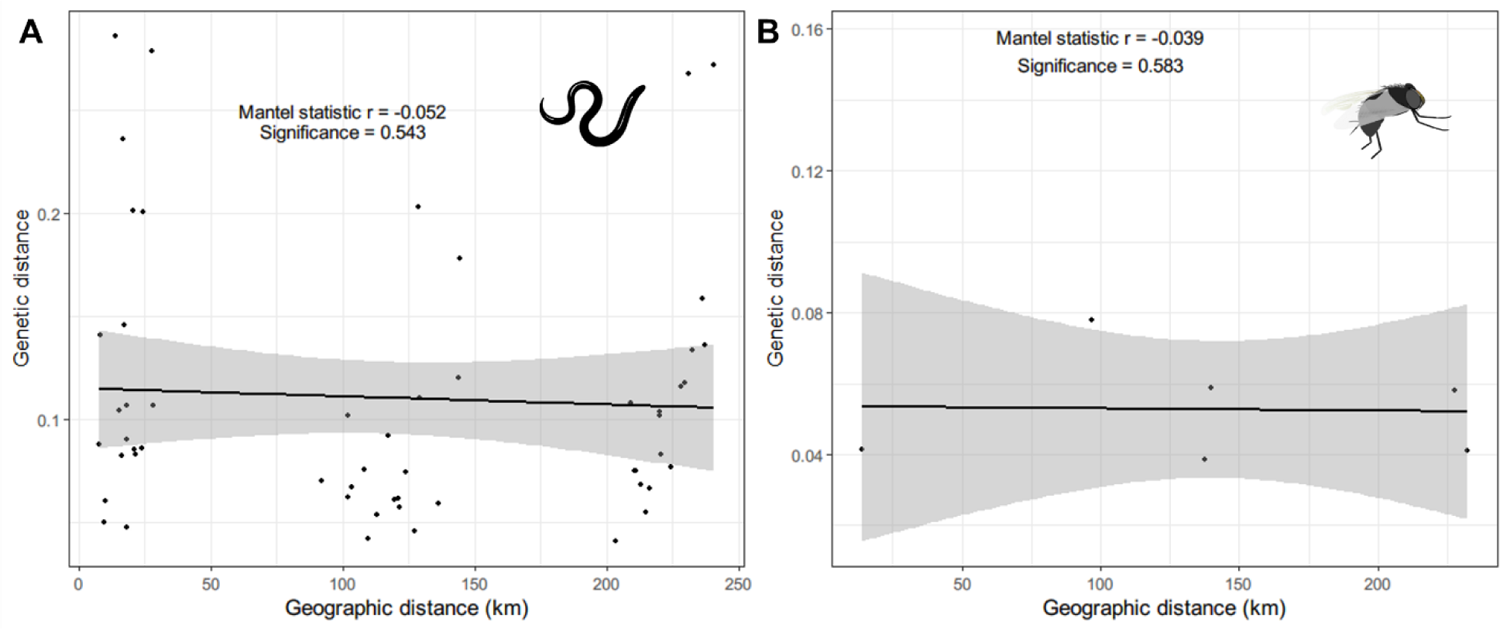
The relationship between the genetic (linearised F_st_) and the Euclidean geographic distances. Isolation-by-distance was tested by the Mantel test, and the significance and the strength of the relationship are shown for the parasite (**A**) and vector (**B**).

### Resistance surface optimisation and testing

We selected five environmental variables for the resistance surface optimisation: elevation, isothermality, soil moisture, flow accumulation and annual precipitation. The values in the resistance surface represent the amount by which the movement is restrained by the given environmental variables. The ecological cost distances obtained for the respective resistance surfaces were used to determine whether the environmental variables could explain the genetic differentiation among parasite and vector sampling locations and performed four replicates of optimisation for 1000 iterations each, then chose the surface with the highest significance (i.e., lowest p-value). For the parasites, we found that the inverse ricker transformation for elevation (r = 0.793, p = 0.005) and soil moisture (r = 0.507, β = 0.002, p = 0.022) were significant (Table 1). The inverse reverse monomolecular transformations for elevation soil moisture were also significant, but the levels of significance were lower compared to the chosen resistance surfaces. Therefore, inverse ricker transformation surfaces for the elevation and soil moisture were used for the preparation of the composite resistance surface map for the parasite data.

**Table 1.**
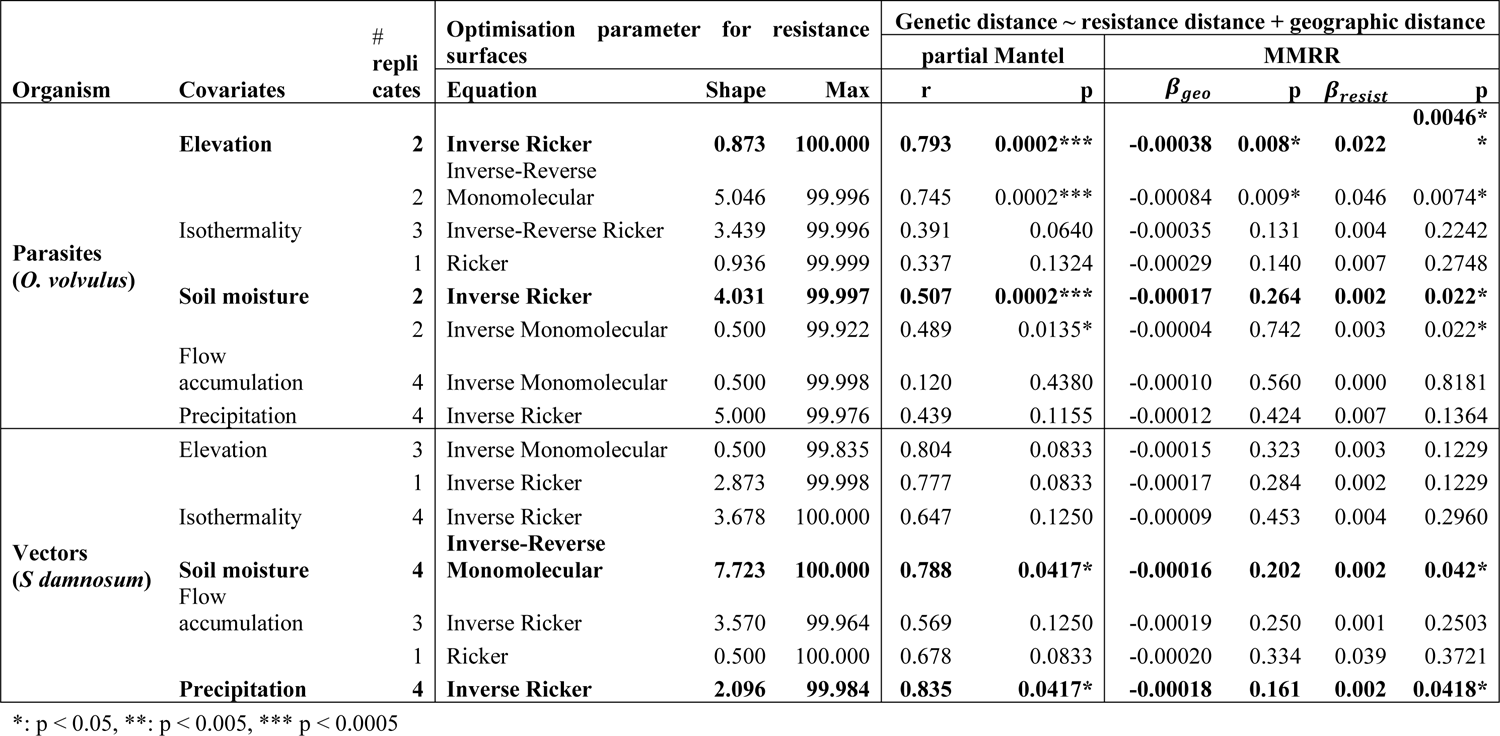
Transformation of environmental surfaces into resistance surfaces with an optimisation function available in *ResistanceGA*. The strength and the direction of association of the resistance surface to the genetic distance are tested with the partial Mantel test and Multiple Matrix Regression with Randomisation (MMRR). The bold transformations are the selected resistance surfaces with the asterisks (*) representing the significance of the coefficients. β_geo_ and β_resist_ represents the regression coefficients for the geographic distance and the cost distance due to the resistance surface respectively.

The inverse ricker transformation was significant in both environmental layers with high resistance to gene flow in the low and high environmental values and lower resistance in the moderate range of environmental values, but with different scale parameters. The resistance to gene flow was lowest (< 30% of the total resistance) in areas with an elevation range of 90– 150 m and in areas with soil moisture of 60–190 mm (Figure 5). A composite resistance surface map was prepared, which showed high resistance around the western parts of the study area, which are characterised by low soil moisture (i.e., Bui National Park in the west, a woodland Savannah zone [96]) and higher elevation. The areas around Lake Volta also have high resistance. Accordingly, the movement corridor map suggests that there is relatively lower connectivity of parasites in the northwestern part of the study area (Figure 6). The central parts of the study area are characterised by high connectivity, showing a potential route for the movement/transmission of parasites.

**Figure 5.**
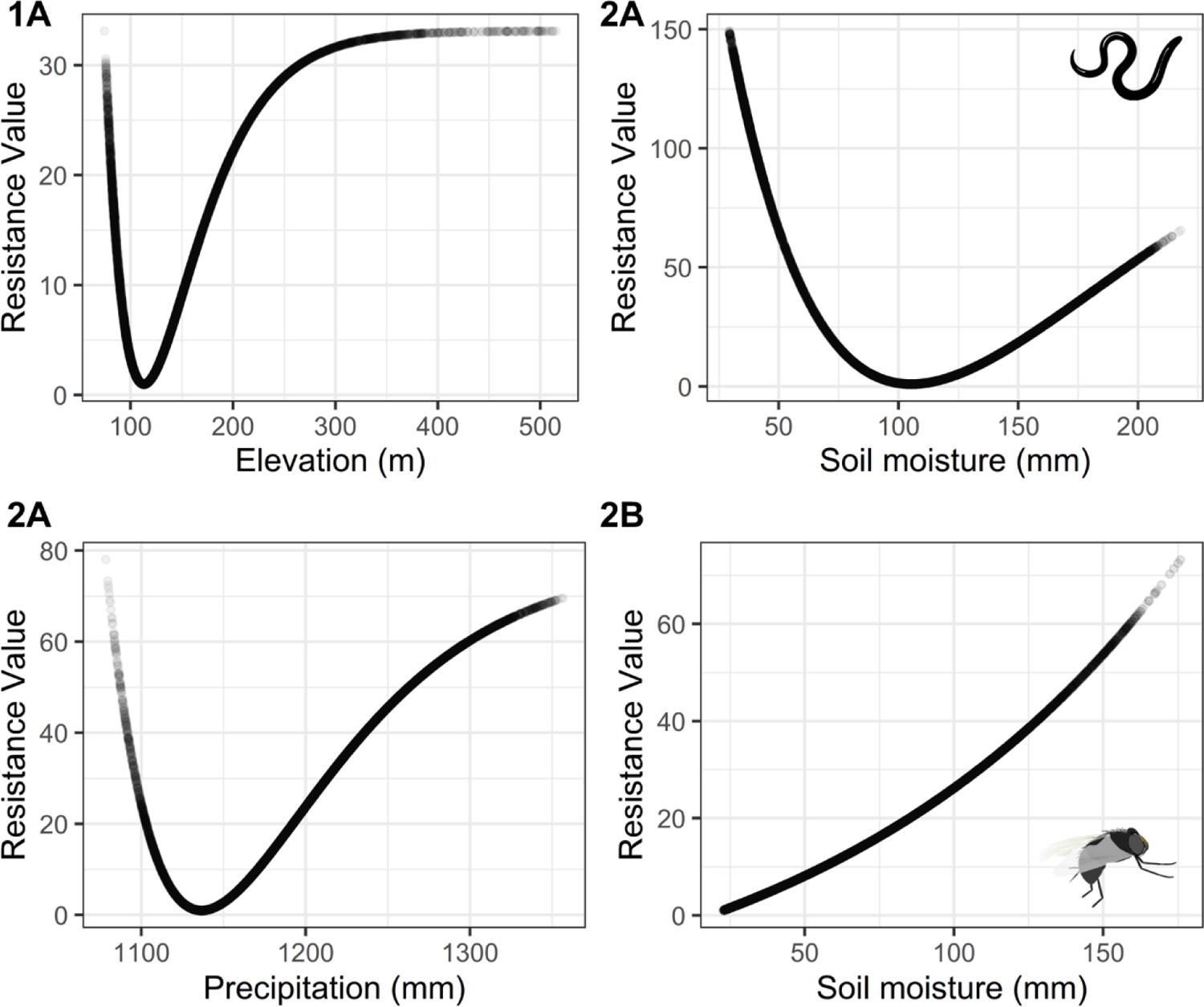
Transformation functions for the significant environmental covariates. The figure shows the relationship between the environmental variables with the resistance against gene flow of the *O. volvulus* (**1A, 1B**) and *S. damnosum* (**2A, 2B**).

**Figure 6.**
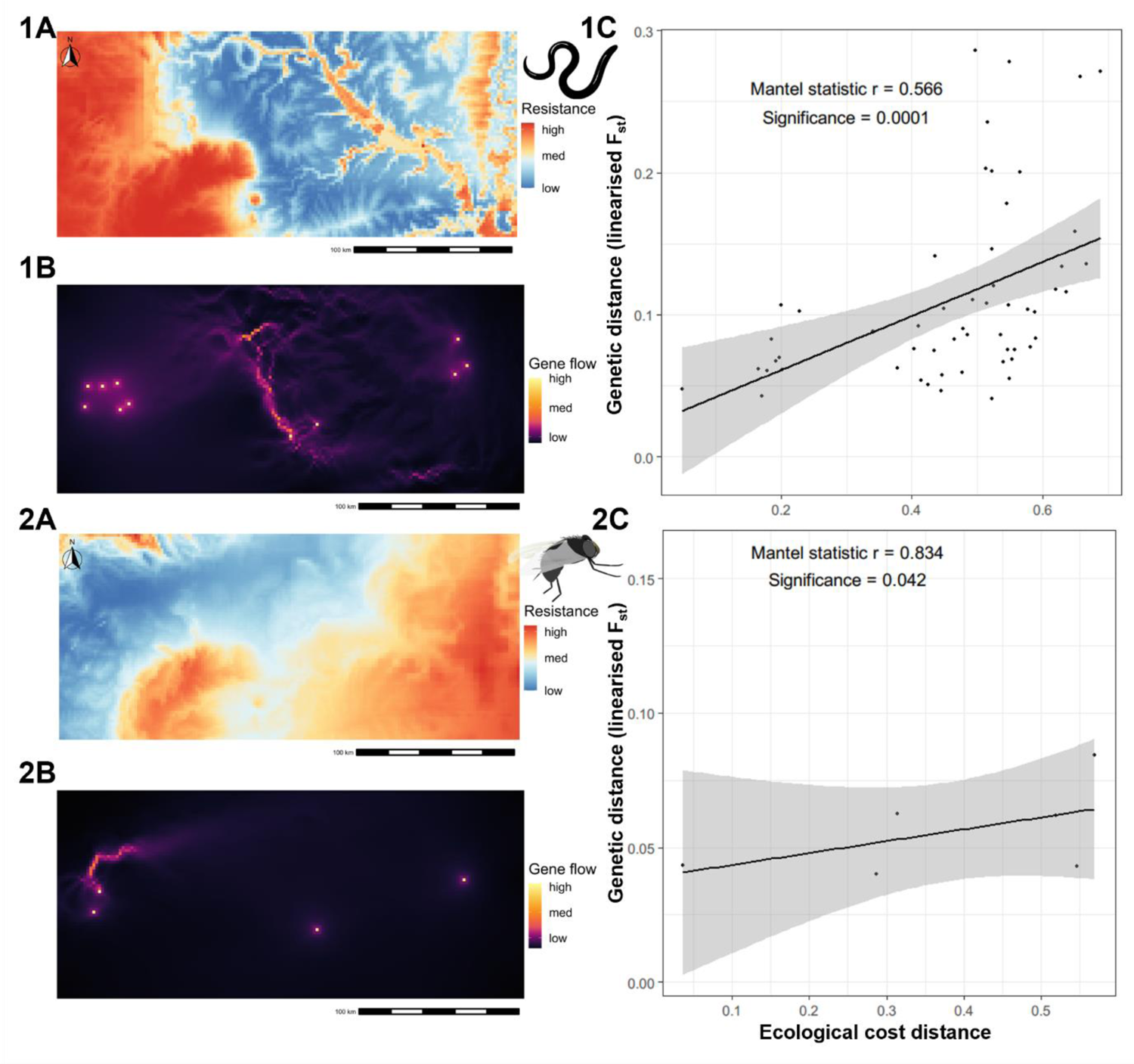
Composite resistance surface maps prepared from the significant environmental variables along with the gene flow map obtained based on the composite resistance surface map and its relationship with the observed genetic distance. The resistance surface maps (1A, 2A) indicate the ease of movement for the parasite and the vector, and the gene flow map (1B, 2B) is obtained based on it with areas highlighted yellow showing the potential routes of movement/gene flow of the organism of interest. The relationship between the ecological distance (the cost distance obtained based on the resistance surface) and the genetic distance (linearised F_st_) (1C, 2C) is shown.

For the vector genetic data, resistance surfaces obtained from soil moisture (r = 0.788, p = 0.0417) and precipitation (r = 0.835, p = 0.0417) were significant, with inverse reverse monomolecular and inverse ricker transformations, respectively. The lowest resistance (< 30% of the maximum resistance) for vector gene flow was in the areas with soil moisture of 22– 90 mm and precipitation of 110–120 cm. These two resistance surfaces were rescaled and merged to create a composite resistance surface as performed on the parasite data. The composite resistance surface for the vectors revealed that there was particularly low resistance for gene flow along the western and northwestern areas of the study area and a moderate level of resistance in the central region. The current density map also showed a higher level of connectivity (lower resistance) around the southwestern Savannah region (Figure 6).

The bivariate map (Figure 7) obtained by combining mf prevalence map and the conductance surface (inverse of resistance surface maps where high conductance implies high suitability for movement) for the parasite shows that the area of high parasite conductance and high prevalence is in the central parts of the transition region of Ghana (Figure 7, Box 2). There is a good correlation between the parasite’s composite conductance surface and the *O. volvulus* infection prevalence map with the majority (57.34%) of sliding window correlation coefficients greater than 0.3 (Figure 7B). Therefore, the areas with high parasite conductance are also the areas of high *O. volvulus* infection prevalence and vice versa. Areas of high vector conductance and high prevalence are found in the central and southwestern parts of the study area. However, a substantial portion of the vector bivariate map has high conductance but low prevalence, particularly around the northwestern region of the study area (Figure 7, Box 1). As a result, the correlation between the conductance map for vectors and the mf infection prevalence is not as strong as the correlation for the parasite counterpart. Only 21.24% of the sliding window correlation coefficients are greater than 0.3 (Figure 7D). There are also the areas in the south-eastern parts of the area that have high prevalence and high parasite conductance; however, low vector conductance (Figure 7, Box 3).

**Figure 7.**
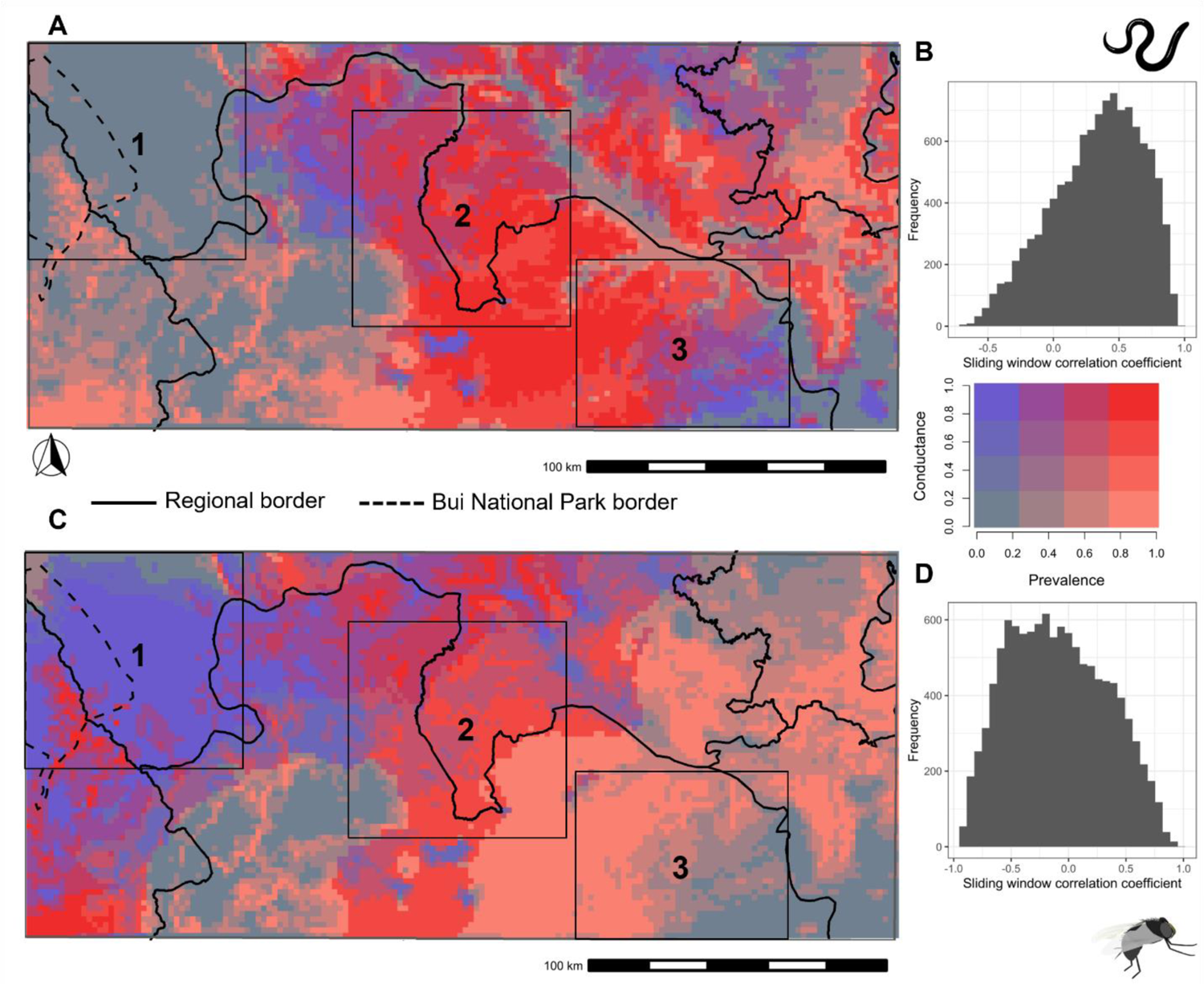
A bivariate map created using composite conductance surfaces and the *Onchocerca volvulus* infection prevalence map. The top row shows the bivariate map for the parasite (**A**) and the bottom row (**C**) for the vector. The legend for the bivariate map is shown on the right, where red colour indicates the areas with high prevalence and high conductance (represents high movement suitability), whereas blue colour indicates areas with high conductance but low prevalence. The histogram on the right of the respective map shows the frequency of the sliding window correlation coefficient between the conductance surface and the prevalence map for the *O. volvulus* infection prevalence map with the parasite (**B**) and the vector (**D**) conductance surface. The solid line represents the regional administrative border, while the broken line shows the border for Bui national park. The three boxes in the figure show contrasting patterns of conductance and prevalence: 1. High vector conductance but low parasite conductance and low *O. volvulus* infection prevalence; 2. High vector and parasite conductance and high *O. volvulus* infection prevalence; 3. Low vector conductance but high parasite conductance and high *O. volvulus* infection prevalence. The conductance and the prevalence on the map are rescaled from 0 to 1.

## Discussion

For the first time in the context of onchocerciasis, we have integrated point location prevalence data, the population genetics of parasites and vectors (as a proxy for parasite and vector movement), and environmental data within a single landscape genetics framework. The visual, spatial representation of parasite and vector movement and infection prevalence shown in Figure 7 is a spatial representation of *O. volvulus* transmission and brings us a step closer to a quantitative, evidence-based method for “delineating” onchocerciasis transmission zones. We have transformed the metrics of genetic connectivity and landscape/ecological variables into a resistance/conductance surface, i.e., a spatial prediction of vector movement and parasite transmission suitability (high resistance or low conductance represents low suitability for movement and transmission and vice versa) which provides an evidence-based methodology by which it may be possible to define transmission zones. For example, geospatially explicit modelling of prevalence and landscape connectivity—can be used to identify reasons for ongoing transmission despite MDAi or newly arisen hot spots of transmission post-MDAi.

Just as the pre-MDAi prevalence is the product of the cumulative history of *O. volvulus* infection, so is the population genetic structure the product of events in the past. Both of these historical elements do not reflect the current transmission patterns of *O. volvulus*. However, using ecological data might enable us to better estimate current transmission as ecological and landscape data are ‘current’. The timeframe over which the climate changes is long compared to the timeframe over which prevalence and population structure changes. Assuming that the ecological parameters are unlikely to have changed significantly over timeframes of either the cumulative infection history giving rise to the prevalence or the microevolutionary processes giving rise to the current population genetic structure, identifying environmental features associated with population genetics and the prevalence allows us to understand current and predict future transmission patterns.

For the ecological transition region of Ghana, the pre-MDAi infection prevalence was positively associated with slope and soil moisture. A likely explanation for this correlation is that greater topological slope results in faster river flow essential for vector breeding. Similarly, soil moisture was also identified to be significant in an analysis of Ethiopian *O. volvulus* nodule prevalence data, where areas with high soil moisture occur in arable land that are usually inhabited by people who are exposed more to vector bites [2,64,97]. In contrast, temperature seasonality was negatively associated with mf prevalence (Additional file Table S4). This is likely because areas with higher fluctuations in temperature might not be favourable for *Simulium*. After all, different species of *Simulium* have different temperature ranges for breeding and biting activities [66], and activities of blood-seeking flies are limited, particularly in low temperatures [98]. Further, the significant relationship between mf prevalence to the temperature seasonality highlights the potential effect of global warming and alterations in annual temperature patterns on the distribution of onchocerciasis. We were not able to detect a significant association of the mf prevalence with the distance to the nearest river, which might be because all the communities surveyed happened to be close to rivers (< 10 km). Therefore, the spatial coverage of the samples might influence the inferred relationship of the ecological variables with the prevalence and the genetic data.

The parasites themselves do not move, however, their movement between geographical locations is mediated either by infected blackflies or infected humans. Population genetics is able to provide insights into the migration of the parasites and the blackflies. The population genetic analyses of parasite and vector genetic data in the ecological transition region of Ghana were largely concordant: both parasite and vector showed low genetic differentiation or high genetic similarity between the sampled communities. Previous studies by Crawford et al. [25] and Gyan [99], suggested the same, i.e., both the parasite and the vector populations were largely genetically homogeneous. Consequently, there was no support for an isolation-by-distance population structure for either the parasites or their vectors in the ecological transition region of Ghana. This suggests that the gene flow of the parasite and the vector populations were not restricted by geographic distance in this study area. However, some degree of genetic differentiation between sampling locations was observed. In order to investigate the likely origins of this relatively weak population structure, we estimated an “ecological distance” parameter from local ecological data for each community, and observed a strong positive correlation. Thus, if “ecological distance” is substituted for “geographical distance” in the isolation-by-distance model, these data do show isolation-by-distance relationships driven by ecological rather than geographical proximity.

With the assumption that environmental factors could explain the resulting observed vector and the parasite genetic connectivity, we used a landscape genetics framework to (1) identify the ecological factors influencing *S. damnosum* and *O. volvulus* population structure then (2) combine the resultant spatial correlation between inferred parasite/vector movement and ecology with the predicted spatial pattern of prevalence to produce an integrated map of likely transmission intensity. Landscape genetics methods combine ecological connectivity with genetic similarity. This allows us to identify the corridors of movement and, thus, the spatially explicit patterns of transmission. It is important to note that high vector connectivity might not necessarily mean high movement suitability, high vector density or high vector biting rates. These are the observed suitability for the movement of blackflies based on the genetic data. High biting rates are crucial for the high endemicity of the disease, whereas vector mobility might help maintain or even amplify onchocerciasis endemicity. Here, we assume that if the vector has high mobility in the areas of high prevalence, there is a likely possibility of high transmission events.

For the parasite population, resistance surfaces obtained from the elevation and soil moisture were significantly associated with the genetic distance. The resistance to parasite gene flow was low (i.e., genetic connectivity was high) in the areas of moderate elevation in the range of 90–150 m and in areas with moderate soil moisture, 60–190 mm. Our estimate of the range of elevation most strongly correlated with prevalence is essentially identical to the range reported proposed by Barro and Oyana [65]. The reason behind high resistance to the parasite gene flow in the areas of low soil moisture could be due to the un-arability of the land and, thus, the lack of human hosts. Soil moisture is reported to be an important environmental feature influencing the occurrence of onchocerciasis in other studies [2, 67]. However, high soil moisture areas might also not be that suitable for onchocerciasis as those were around Lake Volta with non-flowing water and are generally unsuitable for vector breeding. Lake Volta is one of the biggest artificial lakes in the world. Lakes formed by river dams have been reported to affect vector breeding and decrease *O. volvulus* transmission [100–102].

Parasite connectivity indicates where parasite transmission can occur between locations. Blackfly connectivity, in contrast, indicates where transmission may occur between locations due to blackfly movement rather than, or in addition to, human movement. Therefore, differences in the blackfly resistance surface profile compared to the resistance surface for parasites represent the potential transmission mediated by human movement (Figure 6). Further, the blackfly resistance surface was not as strongly correlated as the parasite resistance surface to the mf prevalence map, particularly in the western parts of the study region (Figure 7, Box 1). There are several factors that may contribute to low concordance between blackfly and parasite resistance surfaces. One is the pattern of human population density. For example, the vector connectivity was high in the areas with low soil moisture, while parasite connectivity was low. Low soil moisture indicates lower suitability for agriculture, and they likely have lower human population density and thus appear unsuitable for parasite transmission. A similar case is Bui National Park in the west, where blackflies are present but there is a sparse human settlement and hence low parasite transmission. A second factor is the ratio of *O. volvulus* to *O. ochengi* (and potentially other *Onchocerca* species) in the blackflies. Doyle et al. [103] showed that the proportion of *O. volvulus* larvae in blackflies was lower in western communities compared to the communities in the central and eastern parts of the ecological transition region. The presence of a higher proportion *O. ochengi* has been proposed to impact the vectorial capacity for the *O. volvulus* due to the saturation of the vectors with *O. ochengi* [104, 105].

The weak population structure observed across communities is consistent with the absence of isolation-by-distance observed (Figure 4). The strong correlation between gene flow and several ecological factors related to habitat suitability for black flies indicates that “ecological distance” explains the population genetic structure (Figure 6); i.e., there is a strong correlation between gene flow (genetic differentiation) and ecological connectivity. This strong relationship leads to two important conclusions. First, it provides an explanation for the strong correlation between gene flow and ecological parameters related to blackfly habitat. Second, it suggests a model in which blackfly connectivity is related to the degree to which “local” blackfly populations around discrete breeding sites overlap. What is perhaps surprising is that this proposed overlap between breeding sites extends to create continuous ecological corridors for blackfly movement and parasite transmission.

We produced a bivariate fusion map that combined the results of the mf prevalence and resistance surface mapping (Figure 7). The sliding window correlation coefficient between the surfaces showed a close overlap of the mf prevalence map with the parasite resistance surface, which further validates the landscape genetics output. The bivariate maps represent three different scenarios. Within box 1, there is a high suitability for vector mobility but low infection prevalence and low suitability for parasite mobility. Within box 2, the predicted vector mobility seems to correlate well with parasite mobility and prevalence. In box 3, there is an apparent discordance between the parasite and the vector mobility. The high parasite mobility suggests that the spatial pattern of transmission is likely to be driven more by human movement than vector movement. Therefore, bivariate maps could help in drawing conclusions about what drives transmission in different epidemiological contexts.

Inferences like these might be vital in making spatially explicit onchocerciasis elimination decisions. For example, in the current study, we can hypothesise that communities in the central parts of the study areas (box 2) are one of the critical connecting areas with high suitability for the parasite and the vector gene flow and high onchocerciasis prevalence. The connectivity analysis using the composite resistance surface maps derived from the significant resistance surfaces for the parasites showed that the parasite gene flow was high in the central parts of the ecological transition region of Ghana, around communities from the Bono East (Figure 6). Therefore, MDAi alone might not be sufficient to eliminate onchocerciasis transmission in these areas, where alternative treatment strategies with vector control have to be implemented. However, in areas within box 3, where there is high infection prevalence due to high parasite mobility but low vector mobility, vector control might not be as effective as in the areas within box 2.

Other studies confirm that the communities within box 2 are characterisedparticularly by high biting rates, high vector density and high vector mobility [5, 106] and were among the first to be targeted for both the vector control initially and MDAi later. In addition, this is the area where SOR against ivermectin was first reported [39, 40]. Therefore, with the reports of SOR and the evidence of high gene flow from these areas, the possibility of spreading the SOR strains cannot be ignored. One can expect the consequences of SOR to be spread over an extensive geographical range as a result of the high gene flow of the parasites and the vectors. The approach outlined here might provide an indication of where different epidemiologically relevant phenotypes might likely spread and help design interventions accordingly.

Eliminating onchocerciasis transmission in areas of high connectivity might facilitate onchocerciasis elimination in surrounding areas of lower connectivity. However, it is not to say that the other areas might not act as the source of infection, particularly if the infection is well controlled in the high connectivity region. For example, recent modelling work suggests that low endemic areas can act as a source to re-initiate transmission in MDAi-controlled onchocerciasis endemic areas [107, 108]. Nevertheless, resistance surfaces and connectivity maps could be used to develop heterogeneous intervention strategies to address spatially heterogeneous transmission. Specifically, interventions should be coordinated across locations that are shown to be connected. The intensity of intervention should be varied according to connectivity so that locations of high connectivity receive more intensive interventions than regions of lower connectivity. The rationale is that transmission will be suppressed in a more coordinated fashion with less risk of hotspots of residual transmission even though initial prevalence and transmission may have been highly heterogeneous.

There are some caveats to the current study. First, the sampling density and spatial coverage of the samples in this study are low, and increasing sampling density, in particular, would increase the accuracy of the estimated resistance surfaces. Future landscape genetic studies should consider dense and stratified uniform sampling across space and environmental gradients [29, 109]. Second, due to the unavailability of the nuclear genome sequence data, the genetic analyses utilised mitochondrial sequence data, which might underestimate gene flow [27], and we recommend using nuclear data in future landscape genetics studies. Nevertheless, this study serves as an important use case of the approach with the best data available. Third, the vector resistance surface maps we obtain with the current approach might not necessarily correspond with vector density or vector biting rates. Therefore, incorporating vector abundance data and annual biting rates might further enrich the insights from the approach. Nevertheless, this could be a powerful approach to spatially transforming population genetic connectivity estimates, accounting for ecological variables and predicting routes and geographical boundaries of transmission. Applying this approach to other geographic regions (such as persistent hotspots, cross-border transmission settings and others), and also to other filarial diseases, such as lymphatic filariasis, might prove valuable to the elimination endgame.

## Conclusion

To meet onchocerciasis elimination goals, it is necessary to identify the areas that require intervention via “elimination mapping” (extending prevalence mapping to currently unmapped areas of unknown but probably low prevalence) and by better understanding the spatial patterns of transmission (delineation of transmission zones). We have shown previously how incomplete point prevalence data can be combined with ecological data to provide accurate, spatially continuous, predictions of prevalence [2]. Here we extend that work to provide a novel and promising approach to combine ecological parameters related to vector habitat with population genetic estimates of the vector and the parasite gene flow to produce spatial maps of movement suitability that identify the corridors of movement and give us insight into *O. volvulus* transmission. We demonstrated that the entire ecological transition zone was connected by corridors that are ecologically suitable for vector movement and hence parasite transmission. This leads to the conclusion that the entire ecological transition zone through which the Volta River flows should be treated as a single *O. volvulus* transmission zone. We conclude further that the persistence of transmission across this region, particularly in communities located in the central part of the region, is in part due to the high degree of transmission connectivity over large geographic distances via the “connectivity corridors” we have identified. The spatial pattern of transmission we describe suggests that interventions to interrupt transmission of *O. volvulus* in central Ghana must be coordinated over a large geographical area, particularly decisions to stop MDAi in communities in which local transmission may have been interrupted but which will be subject to re-invasion from surrounding areas in which transmission is yet to be suppressed. We also suggest that landscape genetics could be applied to other vector-borne diseases, particularly lymphatic filariasis, where instances of recrudescence following stop-MDA decisions are accumulating.

## Availability of data and materials

The parasite sequence data are available at NCBI (https://www.ncbi.nlm.nih.gov/ Accession #: PRJNA560089), and the blackfly sequence data have been uploaded to EMBL-EBI (https://www.ebi.ac.uk/ Accession #: PRJEB57094). The onchocerciasis prevalence data were obtained from the ESPEN data portal (https://espen.afro.who.int/tools-resources/download-data), and the sources for the environmental data are provided in the supplementary information. The scripts for the analysis pipeline are uploaded to the GitHub repository (https://github.com/himal2007/landscape_genetics_ghana).

## Abbreviations

BCI: Bayesian credible interval

BIC: Bayesian information criteria

DAPC: Discriminant analysis of principal components

DNA: Deoxyribonucleic Acid

EBI: European Bioinformatics Institute

EMBL: European Molecular Biology Laboratory

ENA: European Nucleotide Archive

ESPEN: Expanded Special Project for Elimination of Neglected Tropical Disease

INLA: Integrated nested Laplace approximations

MDAi: Mass drug administration with ivermectin

MLPE: Maximum likelihood population effects

MMRR: Mixed matrix regression with randomisation

NCBI: National centre for biotechnology information

OCP: Onchocerciasis Control Programme

PCA: Principal component analysis

SD: Standard deviation

SE: Standard error

SNP: Single nucleotide polymorphism

SOR: Sub-optimal response

## Data Availability

The parasite sequence data are available at NCBI (Accession #: PRJNA560089), and the blackfly sequence data have been uploaded to EMBL-EBI (Accession #: PRJEB57094). The onchocerciasis prevalence data were obtained from the ESPEN data portal, and the sources for the environmental data are provided in the supplementary information. The scripts for the analysis pipeline are uploaded to the GitHub repository.

https://github.com/himal2007/landscape_genetics_ghana

https://espen.afro.who.int/tools-resources/download-data

https://www.ebi.ac.uk/

https://www.ncbi.nlm.nih.gov/

## Acknowledgements

The authors wish to acknowledge Anusha Kode for assistance with the sequencing experiment and Dr Kwadwo Frempong for discussion with the results of the prevalence mapping.

## Contributions

Conceptualisation: Himal Shrestha, Shannon M. Hedtke, Warwick N. Grant Data Curation: Himal Shrestha

Formal Analysis: Himal Shrestha

Funding Acquisition: Shannon M. Hedtke, Warwick N. Grant

Investigation: Neha Sirwani, Katie E Crawford, Samuel Armoo, Francis Vierigh Methodology: Himal Shrestha, Shannon M. Hedtke, Karen McCulloch, Rebecca Chisholm Project Administration: Warwick N. Grant, Shannon M. Hedtke

Resources: Samuel Armoo, Francis Vierigh

Supervision: Shannon M. Hedtke, Warwick N. Grant, Rebecca Chisholm, Mike Osei-Atweneboana

Validation: Himal Shrestha, Karen McCulloch, Rebecca Chisholm, Warwick N. Grant, Shannon M. Hedtke

Visualisation: Himal Shrestha

Writing – Original Draft Preparation: Himal Shrestha

Writing – Review & Editing: Himal Shrestha, Shannon M. Hedtke, Warwick N. Grant, Karen McCulloch, Samuel Armoo, Rebecca Chisholm, Katie Crawford

