## Additional file for "Synthesising environmental, epidemiological, and genetic data to assist decision making for onchocerciasis elimination"

**Table S1. Sequencing metadata for each blackfly sample including the communities and the sampling dates.** WGS: Whole Genome Sequencing

| **Sequencing ID** | **Sample ID** | **Sample location: country** | **Community** | **Sample collection date** | **WGS/ amplicon/ etc** | **Library kit** | **Sequencing platform** | **Read length** | **Number of read pairs** | **Trim minimum size** | **Number of trimmed read pairs** | **% pairs retained after trimming** | **Mapped to genome/contig name(s)** | **# reads mapped** | **Number secondary/supplementary [sorted: secondary/suppl]** | **# reads mapped (no suppl)** | **Number mapped (unique)** | **% duplication [metrics]** | **Number uniquely mapped to mitochondrial** | **Average depth mitochondrial** | **Coverage 5x mitochondrial** | **Coverage 20x mitochondrial** |
| --- | --- | --- | --- | --- | --- | --- | --- | --- | --- | --- | --- | --- | --- | --- | --- | --- | --- | --- | --- | --- | --- | --- |
| Sim_GHA_AGB01_HTM3YDRXY_AGTGTTGCAC-CGTGTACCAG_L001 | SIM_GHA_AGB01 | Ghana | Agbelekame | 23-Aug-13 | WGS | Illumina DNA | NovaSeq | 150 | 4152363 | 125 | 3129370 | 75.36 | SIM_GHA_OH30_REF (mt) | 136709 | 6936 | 129773 | 88170 | 32.02% | 88170 | 806.53 | 99.96 | 99.95 |
| Sim_GHA_AGB02_HTM3YDRXY_GACACCATGT-TACACGTTGA_L001 | SIM_GHA_AGB02 | Ghana | Agbelekame | 23-Aug-13 | WGS | Illumina DNA | NovaSeq | 150 | 3445366 | 125 | 2473646 | 71.8 | SIM_GHA_OH30_REF (mt) | 30814 | 1811 | 29003 | 24664 | 14.85% | 24664 | 218.31 | 99.96 | 99.95 |
| Sim_GHA_AGB03_HTM3YDRXY_CCTGTCTGTC-TCACAACAGT_L001 | SIM_GHA_AGB03 | Ghana | Agbelekame | 23-Aug-13 | WGS | Illumina DNA | NovaSeq | 150 | 4959961 | 125 | 3815655 | 76.93 | SIM_GHA_OH30_REF (mt) | 68884 | 3319 | 65565 | 52750 | 19.48% | 52750 | 479.18 | 99.67 | 99.64 |
| Sim_GHA_AGB04_HTM3YDRXY_TGATGTAAGA-AAGGACGCAC_L001 | SIM_GHA_AGB04 | Ghana | Agbelekame | 23-Aug-13 | WGS | Illumina DNA | NovaSeq | 150 | 3731252 | 125 | 2755069 | 73.84 | SIM_GHA_OH30_REF (mt) | 99720 | 5831 | 93889 | 70330 | 25.06% | 70330 | 644.95 | 100 | 100 |
| Sim_GHA_AGB05_HTM3YDRXY_GGAATTGTAA-AGGATGTGCT_L001 | SIM_GHA_AGB05 | Ghana | Agbelekame | 23-Aug-13 | WGS | Illumina DNA | NovaSeq | 150 | 2923138 | 125 | 1985299 | 67.92 | SIM_GHA_OH30_REF (mt) | 57408 | 3061 | 54347 | 46226 | 14.89% | 46226 | 422.89 | 100 | 99.98 |
| Sim_GHA_AGB06_HTM3YDRXY_GCATAAGCTT-TGCGACGGAA_L001 | SIM_GHA_AGB06 | Ghana | Agbelekame | 23-Aug-13 | WGS | Illumina DNA | NovaSeq | 150 | 2510360 | 125 | 1472057 | 58.64 | SIM_GHA_OH30_REF (mt) | 21542 | 1037 | 20505 | 18197 | 11.13% | 18197 | 162.54 | 99.95 | 99.93 |
| Sim_GHA_AGB07_HTM3YDRXY_CTGAGGAATA-AGTGGTTAAG_L001 | SIM_GHA_AGB07 | Ghana | Agbelekame | 23-Aug-13 | WGS | Illumina DNA | NovaSeq | 150 | 2450068 | 125 | 1946554 | 79.45 | SIM_GHA_OH30_REF (mt) | 56458 | 1981 | 54477 | 45508 | 16.44% | 45508 | 417.71 | 100 | 99.98 |
| Sim_GHA_AGB08_HTM3YDRXY_AACGCACGAG-TATCCGAGGC_L001 | SIM_GHA_AGB08 | Ghana | Agbelekame | 23-Aug-13 | WGS | Illumina DNA | NovaSeq | 150 | 4122040 | 125 | 3340784 | 81.05 | SIM_GHA_OH30_REF (mt) | 153529 | 8308 | 145221 | 85984 | 40.75% | 85984 | 787.01 | 100 | 100 |
| Sim_GHA_AGB09_HTM3YDRXY_TCTATCCTAA-CCAGTCGACG_L001 | SIM_GHA_AGB09 | Ghana | Agbelekame | 23-Aug-13 | WGS | Illumina DNA | NovaSeq | 150 | 4979985 | 125 | 3972174 | 79.76 | SIM_GHA_OH30_REF (mt) | 83198 | 4799 | 78399 | 56360 | 28.03% | 56360 | 510.4 | 100 | 99.96 |
| Sim_GHA_AGB10_HTM3YDRXY_CTCGCTTCGG-TTGACTAGTA_L001 | SIM_GHA_AGB10 | Ghana | Agbelekame | 23-Aug-13 | WGS | Illumina DNA | NovaSeq | 150 | 6369403 | 125 | 4902473 | 76.97 | SIM_GHA_OH30_REF (mt) | 1672 | 73 | 1599 | 1469 | 7.63% | 1469 | 12.81 | 97.8 | 10.08 |
| Sim_GHA_AGB11_HTM3YDRXY_CTGTTGGTCC-AACGGTCTAT_L001 | SIM_GHA_AGB11 | Ghana | Agbelekame | 23-Aug-13 | WGS | Illumina DNA | NovaSeq | 150 | 4842250 | 125 | 3794363 | 78.36 | SIM_GHA_OH30_REF (mt) | 174926 | 10229 | 164697 | 95901 | 41.72% | 95901 | 869.21 | 99.51 | 99.44 |
| Sim_GHA_AGB12_HTM3YDRXY_TTACCTGGAA-CTGGAACTGT_L001 | SIM_GHA_AGB12 | Ghana | Agbelekame | 23-Aug-13 | WGS | Illumina DNA | NovaSeq | 150 | 5288906 | 125 | 4196478 | 79.34 | SIM_GHA_OH30_REF (mt) | 185263 | 8759 | 176504 | 113632 | 35.56% | 113632 | 1038.64 | 100 | 100 |
| Sim_GHA_AGB13_HTM3YDRXY_TGGCTAATCA-CTACATGCCT_L001 | SIM_GHA_AGB13 | Ghana | Agbelekame | 23-Aug-13 | WGS | Illumina DNA | NovaSeq | 150 | 4119832 | 125 | 3318046 | 80.54 | SIM_GHA_OH30_REF (mt) | 125659 | 5792 | 119867 | 82916 | 30.76% | 82916 | 755.64 | 99.97 | 99.97 |
| Sim_GHA_AGB14_HTM3YDRXY_AACACTGTTA-TGAGACTTGC_L001 | SIM_GHA_AGB14 | Ghana | Agbelekame | 23-Aug-13 | WGS | Illumina DNA | NovaSeq | 150 | 6695321 | 125 | 5275783 | 78.8 | SIM_GHA_OH30_REF (mt) | 192923 | 9027 | 183896 | 104012 | 43.38% | 104012 | 947.75 | 100 | 100 |
| Sim_GHA_AGB15_HTM3YDRXY_ATTGCGCGGT-GCGGAGCCAA_L001 | SIM_GHA_AGB15 | Ghana | Agbelekame | 23-Aug-13 | WGS | Illumina DNA | NovaSeq | 150 | 4004846 | 125 | 3170794 | 79.17 | SIM_GHA_OH30_REF (mt) | 49554 | 1845 | 47709 | 39724 | 16.64% | 39724 | 354.36 | 100 | 100 |
| Sim_GHA_AGB16_HTM3YDRXY_TGGCGCGAAC-AGTATCAGTT_L001 | SIM_GHA_AGB16 | Ghana | Agbelekame | 23-Aug-13 | WGS | Illumina DNA | NovaSeq | 150 | 4282188 | 125 | 3389487 | 79.15 | SIM_GHA_OH30_REF (mt) | 288624 | 12355 | 276269 | 146272 | 47.03% | 146272 | 1350.74 | 100 | 100 |
| Sim_GHA_AGB17_HTM3YDRXY_TAATGTGTCT-TATGCCTTAC_L001 | SIM_GHA_AGB17 | Ghana | Agbelekame | 23-Aug-13 | WGS | Illumina DNA | NovaSeq | 150 | 4256105 | 125 | 3302783 | 77.6 | SIM_GHA_OH30_REF (mt) | 18358 | 957 | 17401 | 14786 | 14.36% | 14786 | 113.67 | 99.99 | 99.98 |
| Sim_GHA_AGB19_HTM3YDRXY_AGGATGTGCT-GGAATTGTAA_L001 | SIM_GHA_AGB19 | Ghana | Agbelekame | 23-Aug-13 | WGS | Illumina DNA | NovaSeq | 150 | 4893011 | 125 | 3827833 | 78.23 | SIM_GHA_OH30_REF (mt) | 10621 | 452 | 10169 | 9284 | 7.84% | 9284 | 63.37 | 99.96 | 99.79 |
| Sim_GHA_AGB20_HTM3YDRXY_CACGGAACAA-GTGCTAGGTT_L001 | SIM_GHA_AGB20 | Ghana | Agbelekame | 23-Aug-13 | WGS | Illumina DNA | NovaSeq | 150 | 2714711 | 125 | 2177937 | 80.23 | SIM_GHA_OH30_REF (mt) | 109474 | 4290 | 105184 | 75455 | 28.23% | 75455 | 692.57 | 100 | 100 |
| Sim_GHA_AGB21_HTM3YDRXY_TGGAGTACTT-TCCACACAGA_L001 | SIM_GHA_AGB21 | Ghana | Agbelekame | 23-Aug-13 | WGS | Illumina DNA | NovaSeq | 150 | 6084354 | 125 | 4638628 | 76.24 | SIM_GHA_OH30_REF (mt) | 243605 | 12376 | 231229 | 136533 | 40.90% | 136533 | 1245.28 | 100 | 99.97 |
| Sim_GHA_AGB22_HTM3YDRXY_GTATTGACGT-TTGGAATTCC_L001 | SIM_GHA_AGB22 | Ghana | Agbelekame | 23-Aug-13 | WGS | Illumina DNA | NovaSeq | 150 | 10560870 | 125 | 8519170 | 80.67 | SIM_GHA_OH30_REF (mt) | 322451 | 14354 | 308097 | 175424 | 43.01% | 175424 | 1585.76 | 100 | 100 |
| Sim_GHA_AGB23_HTM3YDRXY_CTTGTACACC-AAGCGCGCTT_L001 | SIM_GHA_AGB23 | Ghana | Agbelekame | 23-Aug-13 | WGS | Illumina DNA | NovaSeq | 150 | 3379771 | 125 | 2776582 | 82.15 | SIM_GHA_OH30_REF (mt) | 87176 | 4106 | 83070 | 62511 | 24.67% | 62511 | 566.67 | 100 | 100 |
| Sim_GHA_AGB24_HTM3YDRXY_ACACAGGTGG-ACAACGCTCA_L001 | SIM_GHA_AGB24 | Ghana | Agbelekame | 23-Aug-13 | WGS | Illumina DNA | NovaSeq | 150 | 4169667 | 125 | 3372726 | 80.89 | SIM_GHA_OH30_REF (mt) | 51270 | 2390 | 48880 | 40526 | 16.87% | 40526 | 356.37 | 100 | 99.99 |
| Sim_GHA_ASU01_HTM3YDRXY_GAACTGAGCG-CGCTCCACGA_L001 | SIM_GHA_ASU01 | Ghana | Asubende | 24-Jun-13 | WGS | Illumina DNA | NovaSeq | 150 | 6179903 | 125 | 4766572 | 77.13 | SIM_GHA_OH30_REF (mt) | 150141 | 8079 | 142062 | 102750 | 27.61% | 102750 | 935.95 | 99.99 | 99.97 |
| Sim_GHA_ASU02_HTM3YDRXY_AGGTCAGATA-TATCTTGTAG_L001 | SIM_GHA_ASU02 | Ghana | Asubende | 24-Jun-13 | WGS | Illumina DNA | NovaSeq | 150 | 1460494 | 125 | 1129092 | 77.31 | SIM_GHA_OH30_REF (mt) | 37148 | 1553 | 35595 | 30487 | 14.29% | 30487 | 278.66 | 100 | 99.99 |
| Sim_GHA_ASU03_HTM3YDRXY_CGTCTCATAT-AGCTACTATA_L001 | SIM_GHA_ASU03 | Ghana | Asubende | 24-Jun-13 | WGS | Illumina DNA | NovaSeq | 150 | 5968654 | 125 | 4623230 | 77.46 | SIM_GHA_OH30_REF (mt) | 150787 | 8533 | 142254 | 92695 | 34.76% | 92695 | 841.75 | 100 | 99.97 |
| Sim_GHA_ASU04_HTM3YDRXY_ATTCCATAAG-CCACCAGGCA_L001 | SIM_GHA_ASU04 | Ghana | Asubende | 24-Jun-13 | WGS | Illumina DNA | NovaSeq | 150 | 3143637 | 125 | 2205284 | 70.15 | SIM_GHA_OH30_REF (mt) | 214215 | 12952 | 201263 | 126881 | 36.92% | 126881 | 1163.86 | 100 | 100 |
| Sim_GHA_ASU05_HTM3YDRXY_GACGAGATTA-AGGATAATGT_L001 | SIM_GHA_ASU05 | Ghana | Asubende | 24-Jun-13 | WGS | Illumina DNA | NovaSeq | 150 | 3806500 | 125 | 2909474 | 76.43 | SIM_GHA_OH30_REF (mt) | 214751 | 11644 | 203107 | 126027 | 37.92% | 126027 | 1156.44 | 99.99 | 99.95 |
| Sim_GHA_ASU06_HTM3YDRXY_AACATCGCGC-ACAAGTGGAC_L001 | SIM_GHA_ASU06 | Ghana | Asubende | 24-Jun-13 | WGS | Illumina DNA | NovaSeq | 150 | 6933641 | 125 | 5423772 | 78.22 | SIM_GHA_OH30_REF (mt) | 233242 | 10396 | 222846 | 139181 | 37.49% | 139181 | 1270.01 | 100 | 100 |
| Sim_GHA_ASU07_HTM3YDRXY_CTAGTGCTCT-TACTGTTCCA_L001 | SIM_GHA_ASU07 | Ghana | Asubende | 24-Jun-13 | WGS | Illumina DNA | NovaSeq | 150 | 5485127 | 125 | 4355903 | 79.41 | SIM_GHA_OH30_REF (mt) | 289703 | 12643 | 277060 | 153450 | 44.58% | 153450 | 1411.82 | 100 | 100 |
| Sim_GHA_ASU08_HTM3YDRXY_GATCAAGGCA-ATTAACAAGG_L001 | SIM_GHA_ASU08 | Ghana | Asubende | 24-Jun-13 | WGS | Illumina DNA | NovaSeq | 150 | 6232276 | 125 | 4911081 | 78.8 | SIM_GHA_OH30_REF (mt) | 196219 | 8932 | 187287 | 121367 | 35.15% | 121367 | 1111.51 | 100 | 100 |
| Sim_GHA_ASU09_HTM3YDRXY_GACTGAGTAG-CACTATCAAC_L001 | SIM_GHA_ASU09 | Ghana | Asubende | 24-Jun-13 | WGS | Illumina DNA | NovaSeq | 150 | 6665088 | 125 | 5084042 | 76.28 | SIM_GHA_OH30_REF (mt) | 223556 | 9678 | 213878 | 129395 | 39.46% | 129395 | 1183.81 | 99.99 | 99.97 |
| Sim_GHA_ASU10_HTM3YDRXY_AGTCAGACGA-TGTCGCTGGT_L001 | SIM_GHA_ASU10 | Ghana | Asubende | 24-Jun-13 | WGS | Illumina DNA | NovaSeq | 150 | 8229038 | 125 | 6275413 | 76.26 | SIM_GHA_OH30_REF (mt) | 200639 | 8212 | 192427 | 119683 | 37.73% | 119683 | 1084.41 | 100 | 100 |
| Sim_GHA_ASU11_HTM3YDRXY_CCGTATGTTC-ACAGTGTATG_L001 | SIM_GHA_ASU11 | Ghana | Asubende | 24-Jun-13 | WGS | Illumina DNA | NovaSeq | 150 | 1770179 | 125 | 1394770 | 78.79 | SIM_GHA_OH30_REF (mt) | 25355 | 1430 | 23925 | 20024 | 16.09% | 20024 | 177.12 | 99.97 | 99.9 |
| Sim_GHA_ASU12_HTM3YDRXY_GAGTCATAGG-AGCGCCACAC_L001 | SIM_GHA_ASU12 | Ghana | Asubende | 24-Jun-13 | WGS | Illumina DNA | NovaSeq | 150 | 4938719 | 125 | 3695925 | 74.84 | SIM_GHA_OH30_REF (mt) | 177956 | 11014 | 166942 | 111427 | 33.20% | 111427 | 1011.08 | 99.99 | 99.95 |
| Sim_GHA_ASU13_HTM3YDRXY_CTTGCCATTA-CCTTCGTGAT_L001 | SIM_GHA_ASU13 | Ghana | Asubende | 24-Jun-13 | WGS | Illumina DNA | NovaSeq | 150 | 6104136 | 125 | 4823134 | 79.01 | SIM_GHA_OH30_REF (mt) | 105127 | 8025 | 97102 | 68014 | 29.85% | 68014 | 611.49 | 100 | 100 |
| Sim_GHA_ASU14_HTM3YDRXY_GAAGCGGCAC-AGTAGAGCCG_L001 | SIM_GHA_ASU14 | Ghana | Asubende | 24-Jun-13 | WGS | Illumina DNA | NovaSeq | 150 | 4342461 | 125 | 3255660 | 74.97 | SIM_GHA_OH30_REF (mt) | 137275 | 11107 | 126168 | 85384 | 32.27% | 85384 | 774.43 | 99.64 | 99.63 |
| Sim_GHA_ASU15_HTM3YDRXY_TCCATTGCCG-TCGTGCATTC_L001 | SIM_GHA_ASU15 | Ghana | Asubende | 24-Jun-13 | WGS | Illumina DNA | NovaSeq | 150 | 5192751 | 125 | 4038373 | 77.77 | SIM_GHA_OH30_REF (mt) | 165807 | 10200 | 155607 | 98597 | 36.58% | 98597 | 891.21 | 100 | 100 |
| Sim_GHA_ASU16_HTM3YDRXY_CGGTTACGGC-CTATAGTCTT_L001 | SIM_GHA_ASU16 | Ghana | Asubende | 24-Jun-13 | WGS | Illumina DNA | NovaSeq | 150 | 5809170 | 125 | 4596756 | 79.13 | SIM_GHA_OH30_REF (mt) | 255144 | 11828 | 243316 | 144225 | 40.70% | 144225 | 1323.25 | 100 | 100 |
| Sim_GHA_ASU17_HTM3YDRXY_GAGAATGGTT-TTGCTGCCGA_L001 | SIM_GHA_ASU17 | Ghana | Asubende | 24-Jun-13 | WGS | Illumina DNA | NovaSeq | 150 | 3084932 | 125 | 2466697 | 79.96 | SIM_GHA_OH30_REF (mt) | 117818 | 6461 | 111357 | 83217 | 25.23% | 83217 | 762.78 | 100 | 100 |
| Sim_GHA_ASU18_HTM3YDRXY_AGAGGCAACC-CCATCATTAG_L001 | SIM_GHA_ASU18 | Ghana | Asubende | 24-Jun-13 | WGS | Illumina DNA | NovaSeq | 150 | 2231221 | 125 | 1700402 | 76.21 | SIM_GHA_OH30_REF (mt) | 99878 | 4698 | 95180 | 70018 | 26.39% | 70018 | 636.61 | 99.58 | 99.45 |
| Sim_GHA_ASU19_HTM3YDRXY_CCATCATTAG-AGAGGCAACC_L001 | SIM_GHA_ASU19 | Ghana | Asubende | 24-Jun-13 | WGS | Illumina DNA | NovaSeq | 150 | 7544087 | 125 | 6109560 | 80.98 | SIM_GHA_OH30_REF (mt) | 381483 | 24163 | 357320 | 209069 | 41.46% | 209069 | 1917.04 | 100 | 99.95 |
| Sim_GHA_ASU20_HTM3YDRXY_GATAGGCCGA-GCCATGTGCG_L001 | SIM_GHA_ASU20 | Ghana | Asubende | 24-Jun-13 | WGS | Illumina DNA | NovaSeq | 150 | 4193754 | 125 | 3361366 | 80.15 | SIM_GHA_OH30_REF (mt) | 137681 | 8432 | 129249 | 93684 | 27.47% | 93684 | 853.35 | 100 | 100 |
| Sim_GHA_ASU21_HTM3YDRXY_ATGGTTGACT-AGGACAGGCC_L001 | SIM_GHA_ASU21 | Ghana | Asubende | 24-Jun-13 | WGS | Illumina DNA | NovaSeq | 150 | 1997799 | 125 | 1617042 | 80.94 | SIM_GHA_OH30_REF (mt) | 28599 | 1258 | 27341 | 24006 | 12.04% | 24006 | 214.99 | 100 | 99.98 |
| Sim_GHA_ASU22_HTM3YDRXY_TATTGCGCTC-CCTAACACAG_L001 | SIM_GHA_ASU22 | Ghana | Asubende | 24-Jun-13 | WGS | Illumina DNA | NovaSeq | 150 | 1809276 | 125 | 1422906 | 78.65 | SIM_GHA_OH30_REF (mt) | 37911 | 1818 | 36093 | 30888 | 14.34% | 30888 | 282.14 | 100 | 100 |
| Sim_GHA_ASU23_HTM3YDRXY_ACGCCTTGTT-ACGTTCCTTA_L001 | SIM_GHA_ASU23 | Ghana | Asubende | 24-Jun-13 | WGS | Illumina DNA | NovaSeq | 150 | 4114881 | 125 | 3373054 | 81.97 | SIM_GHA_OH30_REF (mt) | 74591 | 4776 | 69815 | 51845 | 25.68% | 51845 | 469.44 | 100 | 100 |
| Sim_GHA_ASU24_HTM3YDRXY_TTCTACATAC-TTACAGTTAG_L001 | SIM_GHA_ASU24 | Ghana | Asubende | 24-Jun-13 | WGS | Illumina DNA | NovaSeq | 150 | 2379357 | 125 | 1917118 | 80.57 | SIM_GHA_OH30_REF (mt) | 81882 | 5140 | 76742 | 54551 | 28.86% | 54551 | 499.9 | 100 | 100 |
| Sim_GHA_ASU25_HTM3YDRXY_TCATAGATTG-CACCTTAATC_L001 | SIM_GHA_ASU25 | Ghana | Asubende | 24-Jun-13 | WGS | Illumina DNA | NovaSeq | 150 | 5210289 | 125 | 4122139 | 79.12 | SIM_GHA_OH30_REF (mt) | 83983 | 5750 | 78233 | 55459 | 29.00% | 55459 | 494.57 | 100 | 99.99 |
| Sim_GHA_ASU26_HTM3YDRXY_GTATTCCACC-TTGTCTACAT_L001 | SIM_GHA_ASU26 | Ghana | Asubende | 24-Jun-13 | WGS | Illumina DNA | NovaSeq | 150 | 4775105 | 125 | 3777151 | 79.1 | SIM_GHA_OH30_REF (mt) | 86444 | 6430 | 80014 | 58710 | 26.52% | 58710 | 525.72 | 99.96 | 99.94 |
| Sim_GHA_ASU27_HTM3YDRXY_CCTCCGTCCA-CACCGATGTG_L001 | SIM_GHA_ASU27 | Ghana | Asubende | 24-Jun-13 | WGS | Illumina DNA | NovaSeq | 150 | 4499039 | 125 | 3233529 | 71.87 | SIM_GHA_OH30_REF (mt) | 139594 | 8365 | 131229 | 88143 | 32.75% | 88143 | 799.41 | 99.99 | 99.97 |
| Sim_GHA_FOW01_HTM3YDRXY_AACCATAGAA-CCATCTCGCC_L001 | SIM_GHA_FOW01 | Ghana | Fowman-Banda | 24-Jun-13 | WGS | Illumina DNA | NovaSeq | 150 | 5014581 | 125 | 3920516 | 78.18 | SIM_GHA_OH30_REF (mt) | 142379 | 6416 | 135963 | 91733 | 32.47% | 91733 | 835.37 | 100 | 100 |
| Sim_GHA_FOW02_HTM3YDRXY_GGTTGCGAGG-TTGCTCTATT_L001 | SIM_GHA_FOW02 | Ghana | Fowman-Banda | 24-Jun-13 | WGS | Illumina DNA | NovaSeq | 150 | 7379482 | 125 | 5740955 | 77.8 | SIM_GHA_OH30_REF (mt) | 185551 | 6830 | 178721 | 124632 | 30.21% | 124632 | 1130.82 | 99.99 | 99.99 |
| Sim_GHA_FOW03_HTM3YDRXY_TAAGCATCCA-AATGGATTGA_L001 | SIM_GHA_FOW03 | Ghana | Fowman-Banda | 24-Jun-13 | WGS | Illumina DNA | NovaSeq | 150 | 4818815 | 125 | 3886534 | 80.65 | SIM_GHA_OH30_REF (mt) | 57631 | 2803 | 54828 | 44439 | 18.80% | 44439 | 392.22 | 99.53 | 99.44 |
| Sim_GHA_FOW04_HTM3YDRXY_ACCACGACAT-CCGCATACGA_L001 | SIM_GHA_FOW04 | Ghana | Fowman-Banda | 24-Jun-13 | WGS | Illumina DNA | NovaSeq | 150 | 6203598 | 125 | 4645807 | 74.89 | SIM_GHA_OH30_REF (mt) | 146227 | 6288 | 139939 | 100555 | 28.07% | 100555 | 908.46 | 100 | 100 |
| Sim_GHA_FOW05_HTM3YDRXY_GCCGCACTCT-CGAGGTCGGA_L001 | SIM_GHA_FOW05 | Ghana | Fowman-Banda | 24-Jun-13 | WGS | Illumina DNA | NovaSeq | 150 | 5846038 | 125 | 4626900 | 79.15 | SIM_GHA_OH30_REF (mt) | 273170 | 15663 | 257507 | 160871 | 37.49% | 160871 | 1472.24 | 100 | 100 |
| Sim_GHA_FOW06_HTM3YDRXY_CCACCAGGCA-ATTCCATAAG_L001 | SIM_GHA_FOW06 | Ghana | Fowman-Banda | 24-Jun-13 | WGS | Illumina DNA | NovaSeq | 150 | 2961476 | 125 | 2309367 | 77.98 | SIM_GHA_OH30_REF (mt) | 16758 | 765 | 15993 | 14377 | 9.74% | 14377 | 122.34 | 100 | 99.98 |
| Sim_GHA_FOW07_HTM3YDRXY_GTGACACGCA-GTCCGTAAGC_L001 | SIM_GHA_FOW07 | Ghana | Fowman-Banda | 24-Jun-13 | WGS | Illumina DNA | NovaSeq | 150 | 5322271 | 125 | 4046288 | 76.03 | SIM_GHA_OH30_REF (mt) | 153707 | 8150 | 145557 | 105362 | 27.57% | 105362 | 963.16 | 99.67 | 99.66 |
| Sim_GHA_FOW08_HTM3YDRXY_ACAGTGTATG-CCGTATGTTC_L001 | SIM_GHA_FOW08 | Ghana | Fowman-Banda | 24-Jun-13 | WGS | Illumina DNA | NovaSeq | 150 | 3950319 | 125 | 3061186 | 77.49 | SIM_GHA_OH30_REF (mt) | 104194 | 5334 | 98860 | 74628 | 24.44% | 74628 | 678.84 | 100 | 99.96 |
| Sim_GHA_FOW09_HTM3YDRXY_TGATTATACG-TGTAATCGAC_L001 | SIM_GHA_FOW09 | Ghana | Fowman-Banda | 24-Jun-13 | WGS | Illumina DNA | NovaSeq | 150 | 2470932 | 125 | 1980199 | 80.14 | SIM_GHA_OH30_REF (mt) | 77477 | 3021 | 74456 | 58873 | 20.88% | 58873 | 539.52 | 100 | 100 |
| Sim_GHA_FOW10_HTM3YDRXY_CAGCCGCGTA-CACGGCTAGT_L001 | SIM_GHA_FOW10 | Ghana | Fowman-Banda | 24-Jun-13 | WGS | Illumina DNA | NovaSeq | 150 | 3933590 | 125 | 3163077 | 80.41 | SIM_GHA_OH30_REF (mt) | 63661 | 3212 | 60449 | 48812 | 19.16% | 48812 | 440.34 | 100 | 99.98 |
| Sim_GHA_FOW11_HTM3YDRXY_GGTAACTCGC-TCACCAACTT_L001 | SIM_GHA_FOW11 | Ghana | Fowman-Banda | 24-Jun-13 | WGS | Illumina DNA | NovaSeq | 150 | 6022182 | 125 | 4830142 | 80.21 | SIM_GHA_OH30_REF (mt) | 136691 | 5421 | 131270 | 89699 | 31.57% | 89699 | 811.12 | 99.99 | 99.99 |
| Sim_GHA_FOW12_HTM3YDRXY_ACCGGCCGTA-AATATTGCCA_L001 | SIM_GHA_FOW12 | Ghana | Fowman-Banda | 24-Jun-13 | WGS | Illumina DNA | NovaSeq | 150 | 5896701 | 125 | 4656075 | 78.96 | SIM_GHA_OH30_REF (mt) | 149101 | 5903 | 143198 | 101927 | 28.75% | 101927 | 923.54 | 100 | 100 |
| Sim_GHA_FOW13_HTM3YDRXY_TGTAATCGAC-CGCGGTGATC_L001 | SIM_GHA_FOW13 | Ghana | Fowman-Banda | 24-Jun-13 | WGS | Illumina DNA | NovaSeq | 150 | 3599773 | 125 | 2861714 | 79.5 | SIM_GHA_OH30_REF (mt) | 126006 | 5535 | 120471 | 88171 | 26.76% | 88171 | 805.36 | 100 | 99.98 |
| Sim_GHA_FOW14_HTM3YDRXY_GTGCAGACAG-ACGGATGGTA_L001 | SIM_GHA_FOW14 | Ghana | Fowman-Banda | 24-Jun-13 | WGS | Illumina DNA | NovaSeq | 150 | 3740044 | 125 | 2930236 | 78.35 | SIM_GHA_OH30_REF (mt) | 91229 | 5343 | 85886 | 64117 | 25.25% | 64117 | 577.75 | 100 | 100 |
| Sim_GHA_FOW15_HTM3YDRXY_CAATCGGCTG-TTCCTACAGC_L001 | SIM_GHA_FOW15 | Ghana | Fowman-Banda | 24-Jun-13 | WGS | Illumina DNA | NovaSeq | 150 | 2683811 | 125 | 2120168 | 79 | SIM_GHA_OH30_REF (mt) | 77640 | 3770 | 73870 | 58407 | 20.89% | 58407 | 533.57 | 99.97 | 99.97 |
| Sim_GHA_FOW16_HTM3YDRXY_TATGTAGTCA-CATTAGTGCG_L001 | SIM_GHA_FOW16 | Ghana | Fowman-Banda | 24-Jun-13 | WGS | Illumina DNA | NovaSeq | 150 | 5405252 | 125 | 4323428 | 79.99 | SIM_GHA_OH30_REF (mt) | 189375 | 10833 | 178542 | 120568 | 32.40% | 120568 | 1088.76 | 99.6 | 99.44 |
| Sim_GHA_FOW17_HTM3YDRXY_ACTCGGCAAT-TTCAGTTGTC_L001 | SIM_GHA_FOW17 | Ghana | Fowman-Banda | 24-Jun-13 | WGS | Illumina DNA | NovaSeq | 150 | 5740763 | 125 | 4740408 | 82.57 | SIM_GHA_OH30_REF (mt) | 92522 | 5352 | 87170 | 66305 | 23.81% | 66305 | 598.98 | 100 | 100 |
| Sim_GHA_FOW18_HTM3YDRXY_GTCTAATGGC-CCTGACCACT_L001 | SIM_GHA_FOW18 | Ghana | Fowman-Banda | 24-Jun-13 | WGS | Illumina DNA | NovaSeq | 150 | 4183143 | 125 | 3323860 | 79.46 | SIM_GHA_OH30_REF (mt) | 98795 | 4451 | 94344 | 73102 | 22.46% | 73102 | 664.61 | 99.98 | 99.97 |
| Sim_GHA_FOW19_HTM3YDRXY_CCATCTCGCC-AACCATAGAA_L001 | SIM_GHA_FOW19 | Ghana | Fowman-Banda | 24-Jun-13 | WGS | Illumina DNA | NovaSeq | 150 | 2190655 | 125 | 1729200 | 78.94 | SIM_GHA_OH30_REF (mt) | 1259 | 48 | 1211 | 1140 | 4.71% | 1140 | 7.76 | 63.52 | 4.7 |
| Sim_GHA_FOW20_HTM3YDRXY_CTGCGAGCCA-TGGCCGGATT_L001 | SIM_GHA_FOW20 | Ghana | Fowman-Banda | 24-Jun-13 | WGS | Illumina DNA | NovaSeq | 150 | 5957984 | 125 | 4797186 | 80.52 | SIM_GHA_OH30_REF (mt) | 176932 | 6739 | 170193 | 118446 | 30.36% | 118446 | 1079.2 | 100 | 100 |
| Sim_GHA_FOW21_HTM3YDRXY_CGTTATTCTA-AACCTTATGG_L001 | SIM_GHA_FOW21 | Ghana | Fowman-Banda | 24-Jun-13 | WGS | Illumina DNA | NovaSeq | 150 | 1426064 | 125 | 1086340 | 76.18 | SIM_GHA_OH30_REF (mt) | 26909 | 1357 | 25552 | 22142 | 13.22% | 22142 | 199.06 | 100 | 99.99 |
| Sim_GHA_FOW22_HTM3YDRXY_AGATCCATTA-TGGTAGAGAT_L001 | SIM_GHA_FOW22 | Ghana | Fowman-Banda | 24-Jun-13 | WGS | Illumina DNA | NovaSeq | 150 | 4551604 | 125 | 3645100 | 80.08 | SIM_GHA_OH30_REF (mt) | 81677 | 4163 | 77514 | 59748 | 22.82% | 59748 | 536.45 | 100 | 100 |
| Sim_GHA_FOW23_HTM3YDRXY_GTCCTGGATA-TTCGCCACCG_L001 | SIM_GHA_FOW23 | Ghana | Fowman-Banda | 24-Jun-13 | WGS | Illumina DNA | NovaSeq | 150 | 7016572 | 125 | 5567752 | 79.35 | SIM_GHA_OH30_REF (mt) | 199840 | 8673 | 191167 | 130358 | 31.75% | 130358 | 1177.18 | 99.95 | 99.57 |
| Sim_GHA_FOW24_HTM3YDRXY_CAGTGGCACT-CCTATTGTTA_L001 | SIM_GHA_FOW24 | Ghana | Fowman-Banda | 24-Jun-13 | WGS | Illumina DNA | NovaSeq | 150 | 10256220 | 125 | 8052618 | 78.51 | SIM_GHA_OH30_REF (mt) | 242193 | 15199 | 226994 | 144916 | 36.09% | 144916 | 1305.72 | 99.99 | 99.95 |
| Sim_GHA_WIA01_HTM3YDRXY_CCTGCGGAAC-AGCCTATGAT_L001 | SIM_GHA_WIA01 | Ghana | Wia | 9-Aug-15 | WGS | Illumina DNA | NovaSeq | 150 | 3638527 | 125 | 2962547 | 81.42 | SIM_GHA_OH30_REF (mt) | 6820 | 293 | 6527 | 6037 | 7.38% | 6037 | 54.49 | 99.99 | 99.87 |
| Sim_GHA_WIA02_HTM3YDRXY_TTCATAAGGT-CCTTCTAACA_L001 | SIM_GHA_WIA02 | Ghana | Wia | 9-Aug-15 | WGS | Illumina DNA | NovaSeq | 150 | 7574322 | 125 | 6055733 | 79.95 | SIM_GHA_OH30_REF (mt) | 262988 | 11926 | 251062 | 163855 | 34.68% | 163855 | 1496.59 | 100 | 99.97 |
| Sim_GHA_WIA03_HTM3YDRXY_CTCTGCAGCG-TACATCCATC_L001 | SIM_GHA_WIA03 | Ghana | Wia | 9-Aug-15 | WGS | Illumina DNA | NovaSeq | 150 | 6921063 | 125 | 5323280 | 76.91 | SIM_GHA_OH30_REF (mt) | 620466 | 29874 | 590592 | 312177 | 47.12% | 312177 | 2881.53 | 100 | 100 |
| Sim_GHA_WIA04_HTM3YDRXY_CTGACTCTAC-TGACGGCCGT_L001 | SIM_GHA_WIA04 | Ghana | Wia | 9-Aug-15 | WGS | Illumina DNA | NovaSeq | 150 | 7332278 | 125 | 6010038 | 81.97 | SIM_GHA_OH30_REF (mt) | 183843 | 9636 | 174207 | 120972 | 30.49% | 120972 | 1097.77 | 100 | 100 |
| Sim_GHA_WIA05_HTM3YDRXY_TCTGGTATCC-GTAAGCAACG_L001 | SIM_GHA_WIA05 | Ghana | Wia | 9-Aug-15 | WGS | Illumina DNA | NovaSeq | 150 | 9269010 | 125 | 7290944 | 78.66 | SIM_GHA_OH30_REF (mt) | 308821 | 16708 | 292113 | 183410 | 37.16% | 183410 | 1665.48 | 100 | 100 |
| Sim_GHA_WIA06_HTM3YDRXY_CATTAGTGCG-TATGTAGTCA_L001 | SIM_GHA_WIA06 | Ghana | Wia | 9-Aug-15 | WGS | Illumina DNA | NovaSeq | 150 | 13470763 | 125 | 11084288 | 82.28 | SIM_GHA_OH30_REF (mt) | 589509 | 25752 | 563757 | 296361 | 47.38% | 296361 | 2707.45 | 100 | 100 |
| Sim_GHA_WIA07_HTM3YDRXY_ACGGTCAGGA-AACGAGGCCG_L001 | SIM_GHA_WIA07 | Ghana | Wia | 9-Aug-15 | WGS | Illumina DNA | NovaSeq | 150 | 4608412 | 125 | 3456256 | 75 | SIM_GHA_OH30_REF (mt) | 342124 | 15315 | 326809 | 194701 | 40.40% | 194701 | 1798.84 | 100 | 100 |
| Sim_GHA_WIA08_HTM3YDRXY_GGCAAGCCAG-CGGATGCTTG_L001 | SIM_GHA_WIA08 | Ghana | Wia | 9-Aug-15 | WGS | Illumina DNA | NovaSeq | 150 | 2206282 | 125 | 1220142 | 55.3 | SIM_GHA_OH30_REF (mt) | 21593 | 1302 | 20291 | 18227 | 9.99% | 18227 | 163.33 | 100 | 99.99 |
| Sim_GHA_WIA09_HTM3YDRXY_TGTCGCTGGT-AGTCAGACGA_L001 | SIM_GHA_WIA09 | Ghana | Wia | 9-Aug-15 | WGS | Illumina DNA | NovaSeq | 150 | 2837528 | 125 | 2308224 | 81.35 | SIM_GHA_OH30_REF (mt) | 100620 | 6106 | 94514 | 72378 | 23.37% | 72378 | 662.21 | 99.96 | 99.96 |
| Sim_GHA_WIA10_HTM3YDRXY_ACCGTTACAA-TCGCTATGAG_L001 | SIM_GHA_WIA10 | Ghana | Wia | 9-Aug-15 | WGS | Illumina DNA | NovaSeq | 150 | 4144149 | 125 | 3411441 | 82.32 | SIM_GHA_OH30_REF (mt) | 86259 | 3744 | 82515 | 65170 | 20.92% | 65170 | 590.35 | 100 | 100 |
| Sim_GHA_WIA11_HTM3YDRXY_TATGCCTTAC-TAATGTGTCT_L001 | SIM_GHA_WIA11 | Ghana | Wia | 9-Aug-15 | WGS | Illumina DNA | NovaSeq | 150 | 4598322 | 125 | 3795539 | 82.54 | SIM_GHA_OH30_REF (mt) | 114322 | 5521 | 108801 | 83889 | 22.86% | 83889 | 762.81 | 99.95 | 99.95 |
| Sim_GHA_WIA12_HTM3YDRXY_ACAAGTGGAC-AACATCGCGC_L001 | SIM_GHA_WIA12 | Ghana | Wia | 9-Aug-15 | WGS | Illumina DNA | NovaSeq | 150 | 3530308 | 125 | 2544383 | 72.07 | SIM_GHA_OH30_REF (mt) | 116654 | 5920 | 110734 | 82879 | 25.09% | 82879 | 756.45 | 100 | 100 |
| Sim_GHA_WIA13_HTM3YDRXY_TGGTACCTAA-AGTACTCATG_L001 | SIM_GHA_WIA13 | Ghana | Wia | 9-Aug-15 | WGS | Illumina DNA | NovaSeq | 150 | 4499895 | 125 | 3568096 | 79.29 | SIM_GHA_OH30_REF (mt) | 148988 | 7488 | 141500 | 106044 | 25.03% | 106044 | 975.6 | 100 | 99.99 |
| Sim_GHA_WIA14_HTM3YDRXY_TTGGAATTCC-GTATTGACGT_L001 | SIM_GHA_WIA14 | Ghana | Wia | 9-Aug-15 | WGS | Illumina DNA | NovaSeq | 150 | 2767787 | 125 | 2226220 | 80.43 | SIM_GHA_OH30_REF (mt) | 97329 | 4450 | 92879 | 72378 | 22.03% | 72378 | 664 | 100 | 100 |
| Sim_GHA_WIA15_HTM3YDRXY_CCTCTACATG-AGGAGGTATC_L001 | SIM_GHA_WIA15 | Ghana | Wia | 9-Aug-15 | WGS | Illumina DNA | NovaSeq | 150 | 4654383 | 125 | 3731888 | 80.18 | SIM_GHA_OH30_REF (mt) | 54482 | 3168 | 51314 | 42855 | 16.38% | 42855 | 385.56 | 100 | 100 |
| Sim_GHA_WIA16_HTM3YDRXY_GGAGCGTGTA-ACTTACGGAT_L001 | SIM_GHA_WIA16 | Ghana | Wia | 9-Aug-15 | WGS | Illumina DNA | NovaSeq | 150 | 4712028 | 125 | 3581282 | 76 | SIM_GHA_OH30_REF (mt) | 372126 | 13815 | 358311 | 214924 | 40.00% | 214924 | 1981.37 | 99.99 | 99.99 |
| Sim_GHA_WIA17_HTM3YDRXY_GTCCGTAAGC-AAGATACACG_L001 | SIM_GHA_WIA17 | Ghana | Wia | 9-Aug-15 | WGS | Illumina DNA | NovaSeq | 150 | 2678278 | 125 | 1657059 | 61.87 | SIM_GHA_OH30_REF (mt) | 205680 | 9027 | 196653 | 132420 | 32.66% | 132420 | 1229.06 | 100 | 100 |
| Sim_GHA_WIA18_HTM3YDRXY_ACTTCAAGCG-TTCATGGTTC_L001 | SIM_GHA_WIA18 | Ghana | Wia | 9-Aug-15 | WGS | Illumina DNA | NovaSeq | 150 | 3060409 | 125 | 2274243 | 74.31 | SIM_GHA_OH30_REF (mt) | 97778 | 5890 | 91888 | 71199 | 22.46% | 71199 | 647.33 | 99.99 | 99.94 |
| Sim_GHA_WIA19_HTM3YDRXY_TCAGAAGGCG-TATGATGGCC_L001 | SIM_GHA_WIA19 | Ghana | Wia | 9-Aug-15 | WGS | Illumina DNA | NovaSeq | 150 | 9565431 | 125 | 7540488 | 78.83 | SIM_GHA_OH30_REF (mt) | 360200 | 13721 | 346479 | 212472 | 38.64% | 212472 | 1942.18 | 100 | 99.95 |
| Sim_GHA_WIA20_HTM3YDRXY_GCGTTGGTAT-GGAAGTATGT_L001 | SIM_GHA_WIA20 | Ghana | Wia | 9-Aug-15 | WGS | Illumina DNA | NovaSeq | 150 | 5425371 | 125 | 4025314 | 74.19 | SIM_GHA_OH30_REF (mt) | 200516 | 9590 | 190926 | 130208 | 31.77% | 130208 | 1192.33 | 100 | 100 |
| Sim_GHA_WIA21_HTM3YDRXY_ACATATCCAG-ATTGCACATA_L001 | SIM_GHA_WIA21 | Ghana | Wia | 9-Aug-15 | WGS | Illumina DNA | NovaSeq | 150 | 3470836 | 125 | 2656315 | 76.53 | SIM_GHA_OH30_REF (mt) | 121406 | 6940 | 114466 | 86309 | 24.58% | 86309 | 794.15 | 100 | 100 |

**Table S2. The environmental and socio-economic variables that were included in the initial analysis.** The resolution, time period and sources of each of the variables are provided in the table.

| **Categories** | **Covariates** | **Code** | **Resolution** | **Source** | **Time** | **Unit** | **References** |
| --- | --- | --- | --- | --- | --- | --- | --- |
| Temperature | Annual Mean Temperature | BIO1 | 30 arc second (~1 km) | WorldClim V1 Bioclim | 1970-2000 | °C | [1] |
|  | Mean Diurnal Range (Mean of monthly (max temp - min temp)) | BIO2 |  |  |  |  |  |
|  | Temperature Seasonality (standard deviation × 100)^+^ | BIO4 |  |  |  |  |  |
|  | Max Temperature of Warmest Month | BIO5 |  |  |  |  |  |
|  | Min Temperature of Coldest Month^+^ | BIO6 |  |  |  |  |  |
|  | Temperature Annual Range (BIO5-BIO6) | BIO7 |  |  |  |  |  |
|  | Mean Temperature of Wettest Quarter | BIO8 |  |  |  |  |  |
|  | Mean Temperature of Driest Quarter | BIO9 |  |  |  |  |  |
|  | Mean Temperature of Warmest Quarter | BIO10 |  |  |  |  |  |
|  | Mean Temperature of Coldest Quarter | BIO11 |  |  |  |  |  |
|  | Isothermality (BIO2/BIO7) (×100)* | BIO3 |  |  |  | % |  |
|  | Land surface temperature (day) | LSTD |  | MOD11A1.006 Terra Land Surface Temperature and Emissivity Daily Global 1km | 2000-2001, 2010-2012, and 2013-2015 | °C (converted from K) | [2] |
|  | Land surface temperature (night)^+^ | LSTN |  |  |  |  |  |
| Precipitation | Annual Precipitation*^+^ | BIO12 | 30 arc second (~1 km) | WorldClim V1 Bioclim | 1970-2000 | mm | [1] |
|  | Precipitation of Wettest Month | BIO13 |  |  |  |  |  |
|  | Precipitation of Driest Month | BIO14 |  |  |  |  |  |
|  | Precipitation of Wettest Quarter | BIO16 |  |  |  |  |  |
|  | Precipitation of Driest Quarter | BIO17 |  |  |  |  |  |
|  | Precipitation of Warmest Quarter | BIO18 |  |  |  |  |  |
|  | Precipitation of Coldest Quarter | BIO19 |  |  |  |  |  |
|  | Precipitation Seasonality (Coefficient of Variation) | BIO15 |  |  |  | coefficient of variation |  |
| Topographical | Digital Elevation Model* | DEM | 30 arc second (~1 km) | CGIAR-SRTM | 2000 | meters | [3] |
|  | Slope^+^ | SLP |  |  |  | topographic slope in degree |  |
| Vegetation indices | Normalised Difference Vegetation Index | NDVI | 30 arc second (~1 km) | MOD13A2.006 Terra Vegetation Indices 16-Day Global 1km | 2000-2001, 2010-2012, and 2013-2015 | NA | [4] |
|  | Enhanced Vegetation Index | EVI |  |  |  |  |  |
| Hydrological data | Flow accumulation* | FC | 30 arc sec (~1 km) | HydroSHEDS, WWF | 2000 | Number of cells | [5,6] |
|  | Distance to the nearest river (derived from water lines in DIVA-GIS) ^+^ | DW |  |  | 2003 | km | [7] |
|  | Tasseled cap wetness index | TCW | 5 km | Malaria Atlas Project | 2001-2015 | NA | [8] |
|  | Soil moisture*^+^ | SM |  | TerraClimate: Monthly Climate and Climatic Water Balance for Global Terrestrial Surfaces | 2000-2001, 2010-2012, and 2013-2015 | mm | [9] |
| Socio-demographic | Population density (UN - adjusted) | PD | 30 arc second (~1 km) | Gridded Population of World Version 4 (GPWv4) | 2000-2001, 2010-2012, and 2013-2015 | density | [10] |
|  | Improved housing (prevalence) ^+^ | IHP | 30 arc second (~1 km) | Malaria Atlas Project | 2001, 2001-2015, and 2015 | % | [11] |
|  | Night-time lights | NL | 100 m | WorldPop (Resampled DMSP, OLS) | 2000-2001, 2010-2012, and 2013 | NA | [12] |

* Variables selected for the landscape genetics analysis; ^+^ Variables selected for prevalence mapping

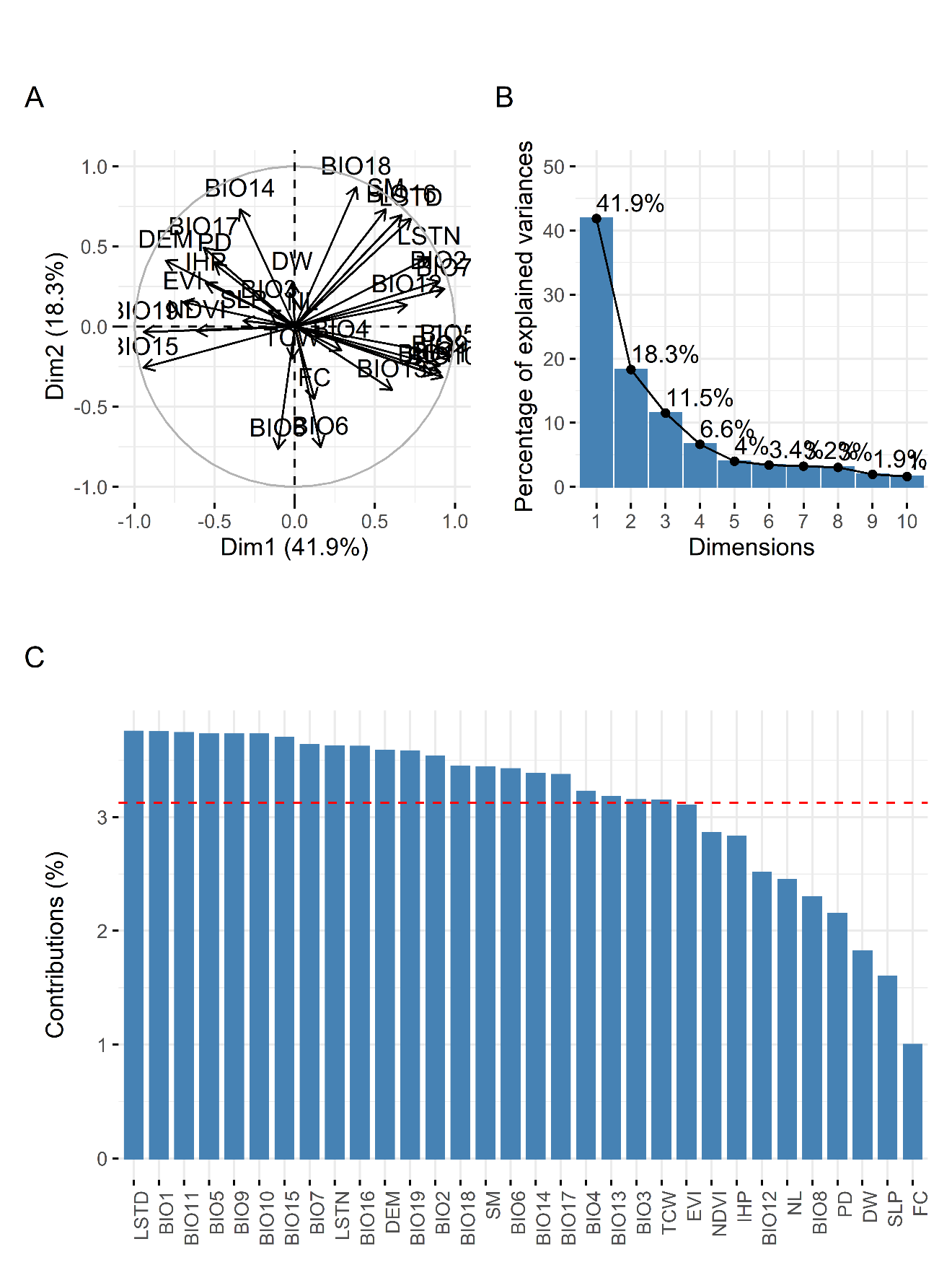

**Figure S1. Principle component analysis on the environmental variables from the locations of prevalence data**. Variable correlation plot (A) showing the extent and the direction of the relationship between variables. Scree plot (B) is showing the percentage of variance explained by the principal components, and the first five dimensions explain > 80% of the total variance. The contribution of the variables in explaining the variability (C) of the first five principal components was used to select the correlated variables.

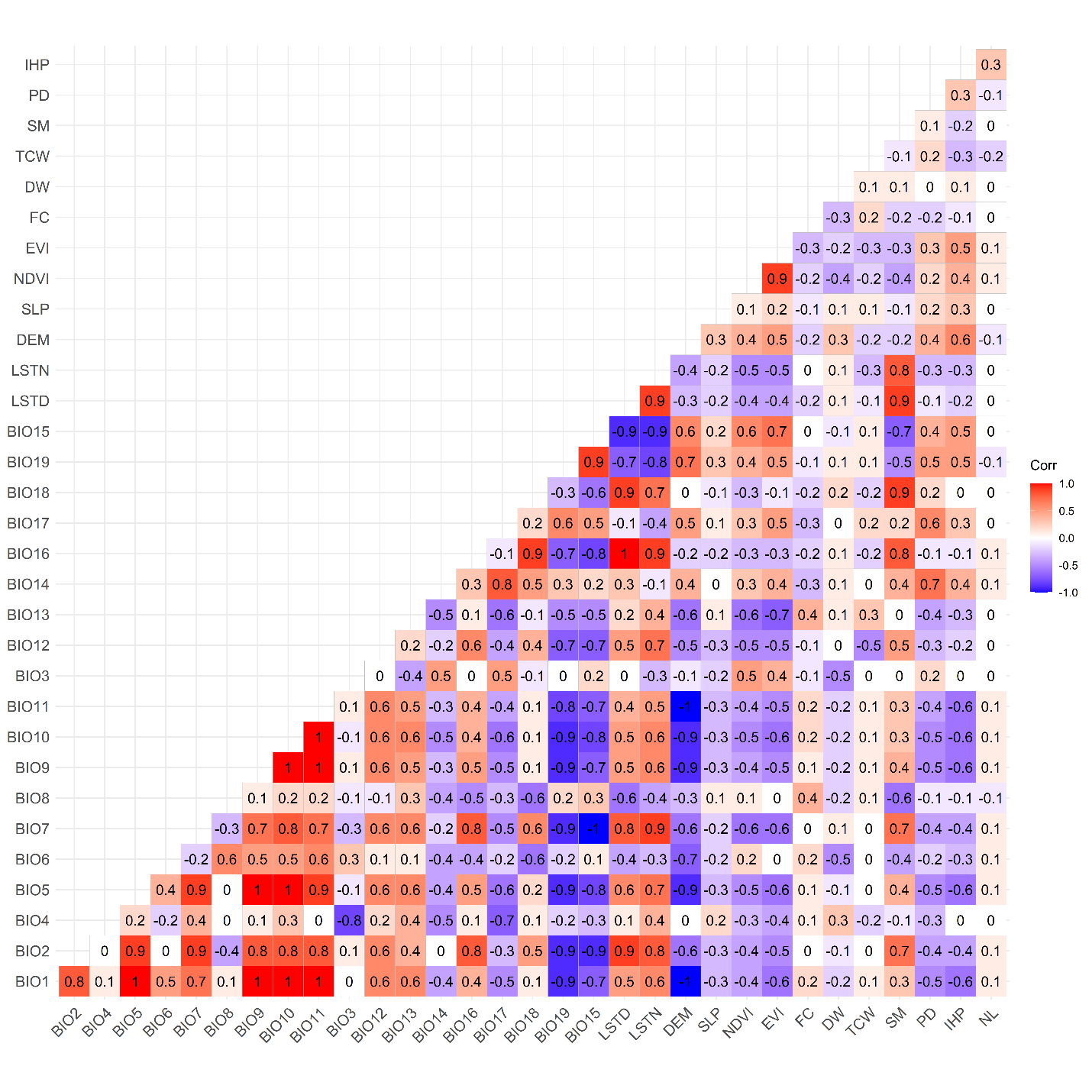
**Figure S2. Correlation matrix between the 32 environmental variables from the sampling locations used for the prevalence data analysis**. The name of the environmental variables is presented in their acronym format.

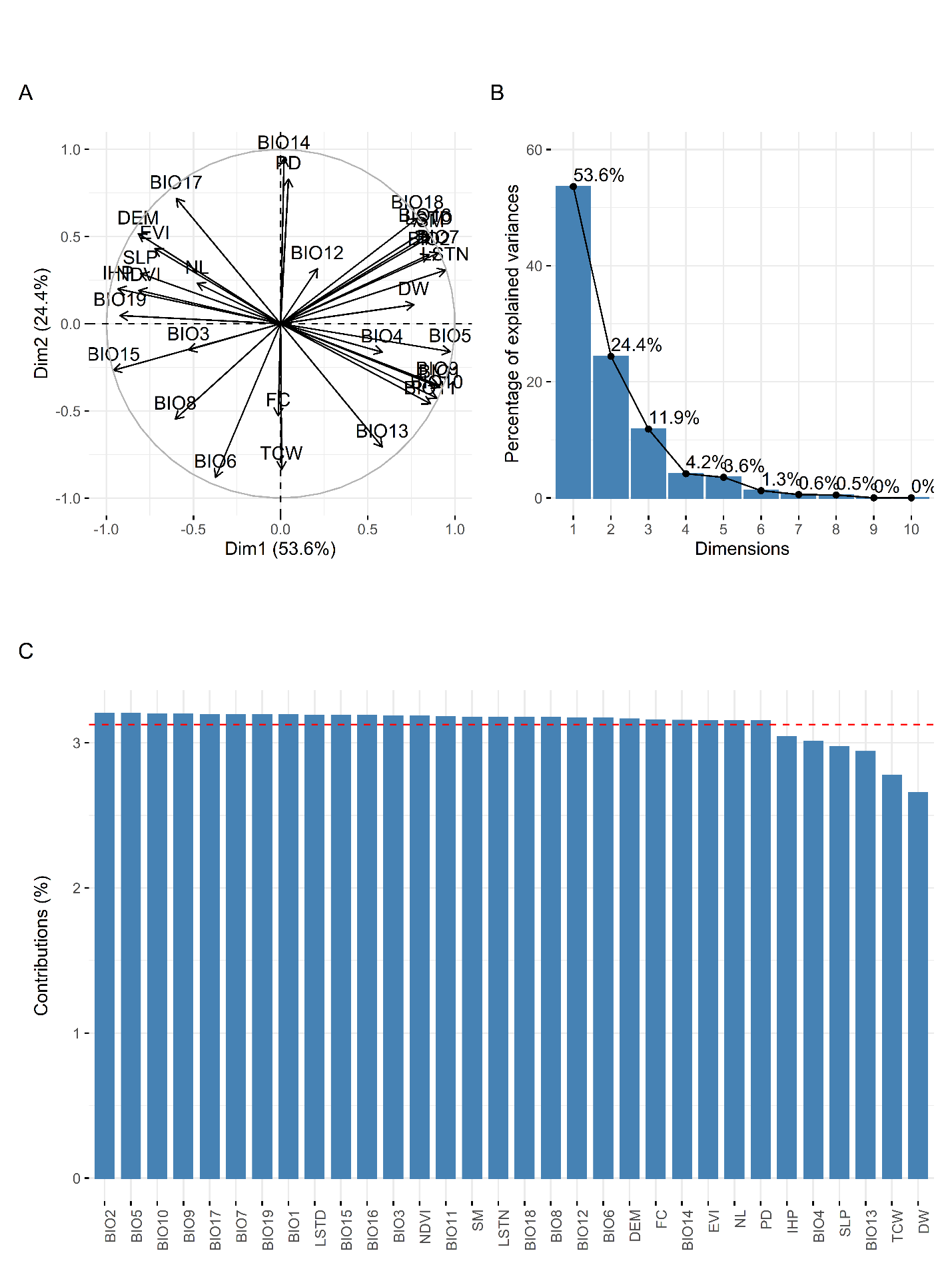

**Figure S3. Principle component analysis on the mean of the environmental variables along the straight path distance between the sampling locations for landscape genetics**. Variable correlation plot (A) showing the extent and the direction of the relationship between variables. Scree plot (B) is showing the percentage of variance explained by the principal components, and the first two dimensions explain >80% of the total variance. The contribution of the variables in explaining the variability (C) of the first five principal components was used to select the correlated variables.

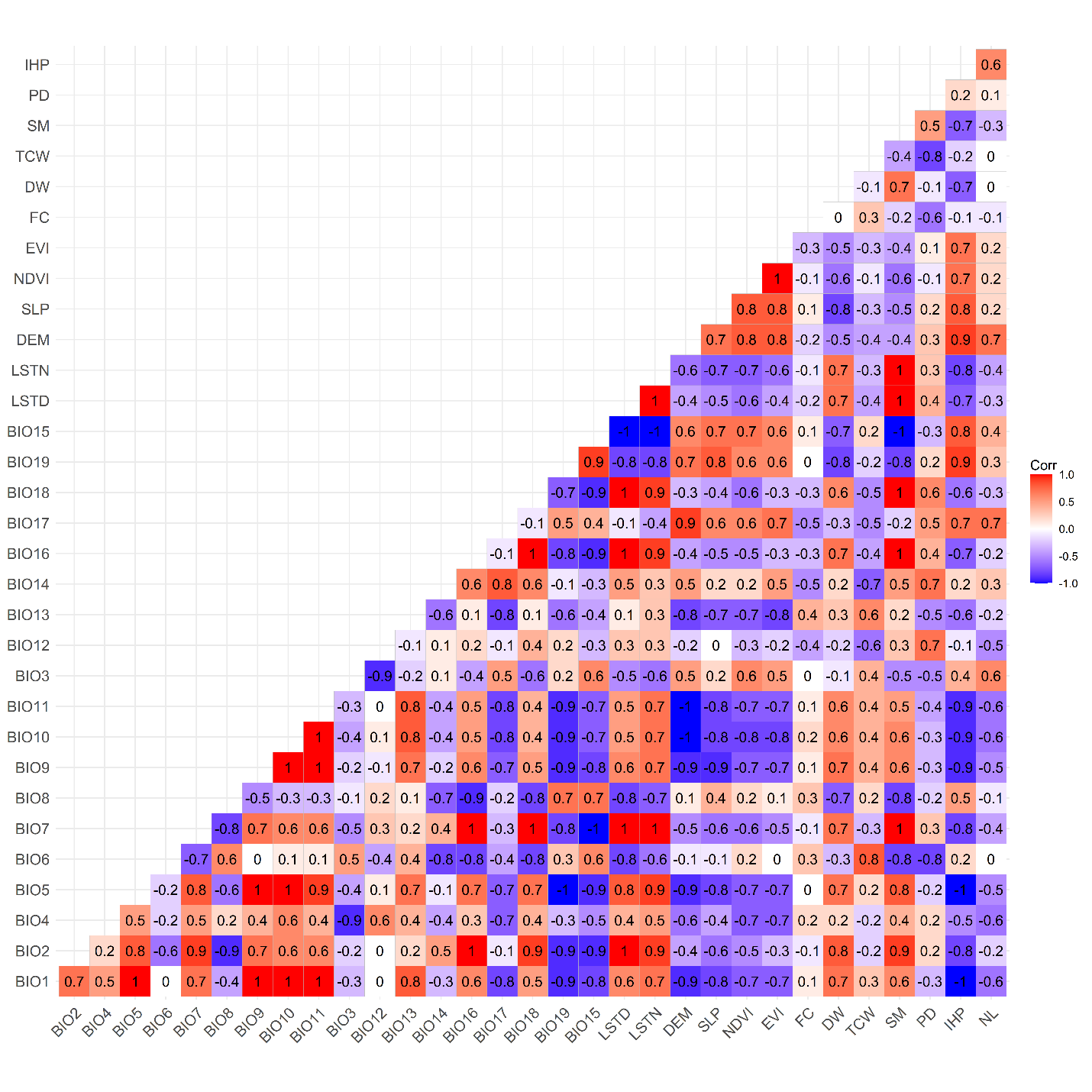

**Figure S4. Correlation matrix between the 32 environmental variables extracted as the mean value encountered along the straight path distance between the sampling locations used for the landscape genetics analysis**. The name of the environmental variables is presented in their acronym format.

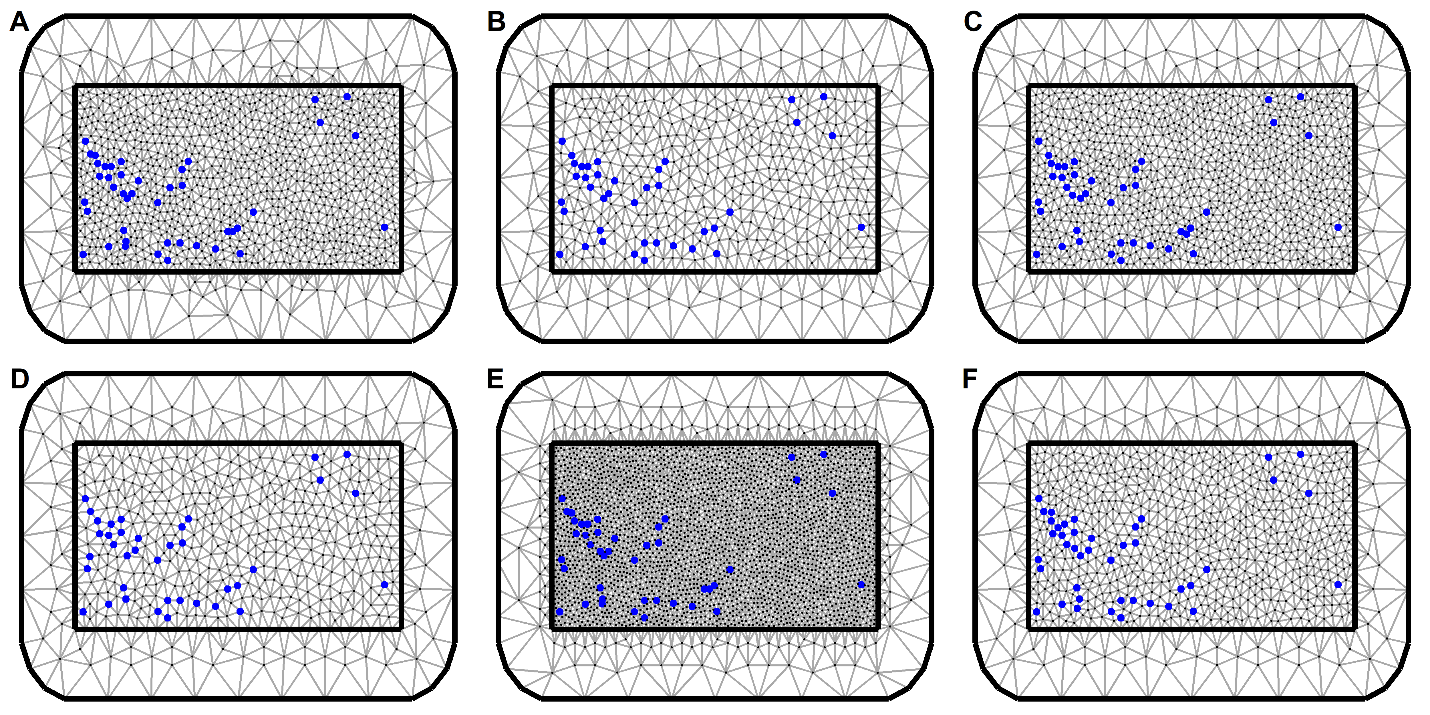

**Figure S5. Different triangulation meshes tested for the model fit and computational cost.** Mesh A was chosen because the trade-off between the model fit, and computation time was optimal compared to other meshes.

**Table S3. Geographic coordinates of the sampling sites along with their site code, government administrative region, sample size and population genetics summary statistics.** Four different sampling locations for the blackflies are shown in the rows below the 15 sampling locations for the parasites.

| **Samples** | **Regions** | **Site name** | **Site code** | **Number of samples (n)** | **Longitude** | **Latitude** | **Number of alleles** | **Mean allelic richness** | **Gene diversity** | **Number of haplotypes** |
| --- | --- | --- | --- | --- | --- | --- | --- | --- | --- | --- |
| Parasites | Savannah | Agborlekame (1) | AB1 | 13 | -2.21 | 8.23 | 227 | 1.0623 | 0.0415 | 13 |
|  |  | Agborlekame (2) | AB2 | 15 | -2.12 | 8.25 | 227 | 1.0579 | 0.0386 | 14 |
|  |  | Wiae Chabbon * | WTC | 14 | -0.21 | 8.28 | 258 | 1.0699 | 0.0466 | 24 |
|  |  | Takumdo* |  | 2 | -0.19 | 8.28 |  |  |  |  |
|  |  | Wiae* |  | 9 | -0.22 | 8.32 |  |  |  |  |
|  | Bono | Bui | BUI | 6 | -2.28 | 8.24 | 216 | 1.0825 | 0.055 | 6 |
|  |  | Kyingakrom | KYG | 15 | -2.11 | 8.1 | 240 | 1.0747 | 0.0498 | 15 |
|  |  | Nyire | NYR | 12 | -2.3 | 8.11 | 236 | 1.0758 | 0.0505 | 12 |
|  | Bono East | New Longoro | NLG | 13 | -2.05 | 8.13 | 236 | 1.079 | 0.0526 | 13 |
|  |  | Ohiampe | OHP | 3 | -1.14 | 7.95 | 201 | 1.0635 | 0.0423 | 3 |
|  |  | Baaya* | BAS | 1 | -1.02 | 8 | 240 | 1.0693 | 0.0462 | 17 |
|  |  | Asubende* |  | 16 | -0.96 | 8.02 |  |  |  |  |
|  |  | Senyase* |  | 2 | -1 | 8.02 |  |  |  |  |
|  | Northern | Jagbengbendo | JAG | 30 | -0.13 | 8.33 | 258 | 1.0637 | 0.0425 | 28 |
|  |  | Kojoboni | KOJ | 13 | -0.18 | 8.49 | 237 | 1.0804 | 0.0536 | 12 |
| Vectors | Savannah | Agborlekame (1) | AGB | 20 | -2.211 | 8.242 | 972 | 1.4964 | 0.1035 | 17 |
|  | Bono | Fawoman-Banda | FOW | 19 | -2.245 | 8.12 | 928 | 1.4388 | 0.0869 | 15 |
|  | Bono East | Asubende | ASU | 26 | -0.981 | 8.017 | 961 | 1.4248 | 0.0981 | 23 |
|  | Northern | Wiae | WIA | 20 | -0.144 | 8.286 | 904 | 1.3905 | 0.0773 | 19 |

* Communities within the geographic distance of 5 km and thus merged and the centroid was taken as the geospatial coordinate for the merged community.

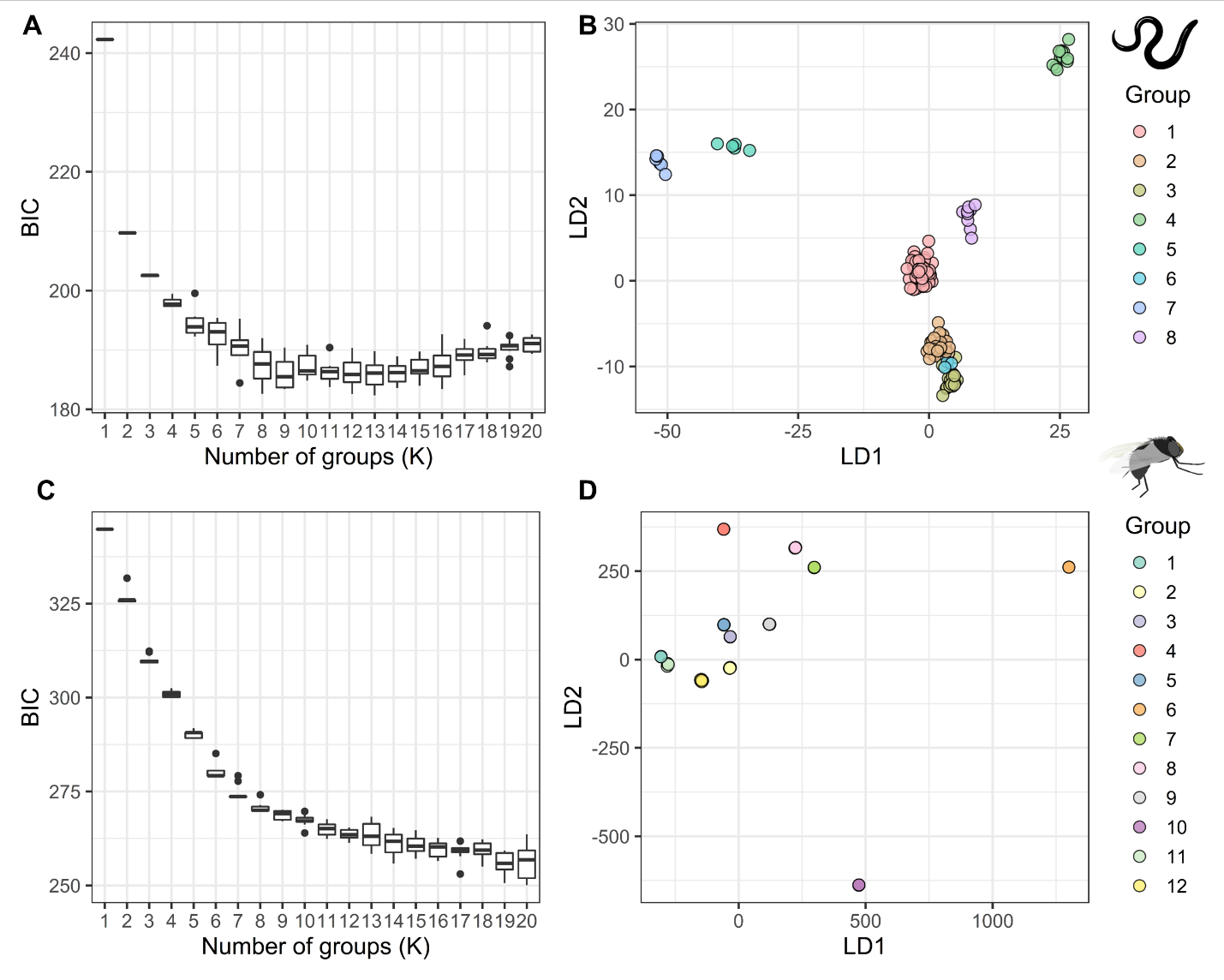

**Figure S6. Clustering analysis on the *O. volvulus* and the *S. damnosum* genetic data.** The optimum number of clusters was determined with the help of BIC scores (**A, C**) and the discriminant analysis of the principal component (DAPC) was done on the inferred clusters (**B, D**) for both the parasite and the vector data.

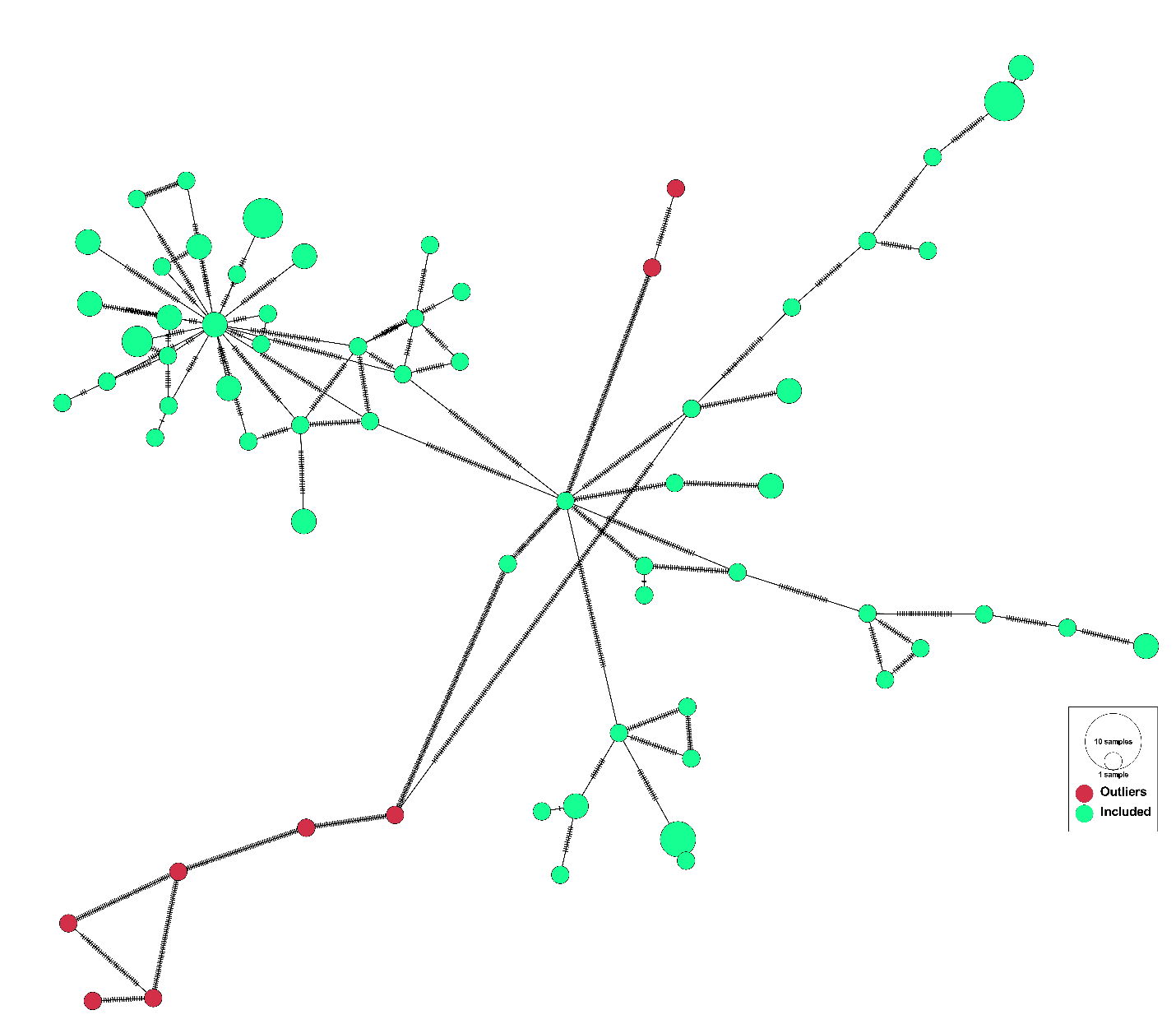

**Figure S7. Haplotype network analysis of the *S. damnosum* samples showing the location of outliers in the haplotype network.** The red nodes are the outlier samples, and the number of hatches between the nodes indicates variation between the nodes. The number of hatches for the edges connecting to outlier nodes is substantially high.

**Table S4. Mean estimates of the posterior regression coefficients for each environmental variable and the hyperparameters while mapping the baseline *O. volvulus* infection prevalence**. The bold environmental covariates are significant based on 95% BCI (Bayesian Credible Interval). Regression coefficients for a particular covariate represent the change in the $logit$ function of prevalence for a unit change in that covariate, given that all other variables are kept constant.

| **Variables** | **Regression coefficients** | |
| --- | --- | --- |
|  | **Mean** | **95% BCI** |
| **Slope** | **2.126** | **(0.032, 4.338)** |
| **Soil moisture** | **0.043** | **(0.004, 0.084)** |
| **Temperature seasonality** | **-0.022** | **(-0.044, -0.001)** |
| Improved housing prevalence | 14.300 | (-31.576, 59.597) |
| Minimum temperature coldest month | 0.249 | (-0.021, 0.535) |
| Distance to the nearest river | 0.174 | (-0.348, 0.701) |
| Annual precipitation | -0.024 | (-0.053, 0.005) |
| Land surface temperature night | -0.777 | (-2.428, 0.807) |
| Intercept | 18.770 | (-36.469, 73.334) |
| **Range** | **4,396.350** | **(1660.26, 7881.19)** |
| **Variance** | **30.905** | **(11.06, 85.812)** |
| 95% BCI includes 0.025 quantiles and the 0.975 quantiles of the posterior probability distribution of the coefficients | | |
